# Oral microbiome profiles relate periodontal disease and brain health - the PAROMIND Study

**DOI:** 10.1101/2025.02.22.25322702

**Authors:** Marvin Petersen, Carolin Walther, Katrin Borof, Guido Heydecke, Thomas Beikler, Malik Alawi, Christian Müller, Felix L. Nägele, Birgit-Christiane Zyriax, Jens Fiehler, Jürgen Gallinat, Simone Kühn, Raphael Twerenbold, Corinna Bang, Götz Thomalla, Bastian Cheng, Ghazal Aarabi

## Abstract

The relationship between oral microbiome composition and brain health in the general population remains poorly understood. In this study, we inferred a microbiome similarity network based on 16S rRNA sequencing data of crevicular fluid collected from 1,026 participants in the Hamburg City Health Study, which revealed a continuous disease gradient mirroring the microbial pathogenicity spectrum of periodontitis. Leveraging this network, we systematically examined associations between periodontal microbiome profiles and 37 brain health-related phenotypes, including cognitive function, brain structure, mental health, inflammatory biomarkers, diet, vascular risk factors, and demographics. Higher abundance of periodontitis-related microbial taxa was linked to poorer cognitive performance, elevated leukocyte counts and lower MIND diet adherence after covariate adjustment, but no significant associations were found for the remaining brain health phenotypes. Notably, we identified both previously known as well as novel microbial associations with brain health phenotypes. These findings advance the understanding of the oral microbiome-brain axis, highlighting potential pathways connecting periodontal health and brain function with potential implications for future causal and interventional studies.

## Introduction

The human oral microbiome harbors a diverse community of microorganisms that influences the health and well-being of their hosts.^1^ In recent years, periodontitis, which is the inflammatory disruption in the host–microbial homeostasis of the periodontal pocket, has gained increasing attention as a key factor impacting brain health. Studies indicate that periodontitis and the linked bacterial communities are associated with the incidence of cognitive decline and Alzheimer’s dementia.^2–7^ Given its estimated prevalence between 10% and 50% in elderly people, this renders periodontitis a relevant public health concern.^8^ As therapeutic interventions can alter the progression of periodontitis, comprehending its impact on the brain is vital for effective prevention and management of cognitive sequelae.

Among bacterial species of the subgingival biofilm, several are known to be associated with periodontitis including: *Porphyromonas gingivalis*, *Tannerella forsythia* and *Treponema denticola*.^9,10^ Mechanistic models have been proposed to explain the connection between these bacterial communities and cognitive decline. The connection is considered to arise from bacterial species promoting systemic inflammation, neurodegenerative processes, and blood-brain barrier disruption.^11–14^

Despite existing research efforts, our understanding of the association between oral microbiome composition and brain health remains limited. While recent studies have predominantly focused on clinical Alzheimer’s disease and mild cognitive impairment cohorts,^7,15–19^ understanding these connections in the general population requires research within broader community-based settings, which remains relatively limited.^20^ Additionally, many investigations are constrained by small sample sizes and a lack of multi-modal imaging and biomarker data potentially leading to inconsistent findings.^7^ These issues are further complicated by the inherent complexity of microbiome data.

Tapping into these research needs, we aim to advance the understanding of the oral microbiome-brain axis by applying advanced analysis techniques capable of integrating the large-scale, multi-domain data crucial for achieving robust insights. Specifically, our primary hypothesis was that subgingival microbial profiles indicative of periodontitis would be associated with variations in brain health-related host phenotypes within a population-based cohort. To investigate this hypothesis, we analyzed population-based data of n=1,026 individuals from the Hamburg City Health Study.^21^ Our approach focuses on a topology-based analysis that integrates abundance data of the subgingival microbiome derived from 16S rRNA sequencing of subgingival samples with in-depth clinical and lifestyle data including oral health assessments, cognitive test scores, mental health scores, neuroimaging, circulating inflammatory markers, dietary patterns, and vascular risk measures.

## Results

Our methodology, which integrates microbiome, clinical and lifestyle data into a unified analysis framework, is illustrated in *figure 1*. Quality assessment of subgingival microbiome abundance and brain MR imaging data resulted in a final analysis sample of 1,026 individuals ([mean ± SD] age 63.72 ± 8.2 years, 42.7% female; for details see *table 1*).

**Figure 1.**
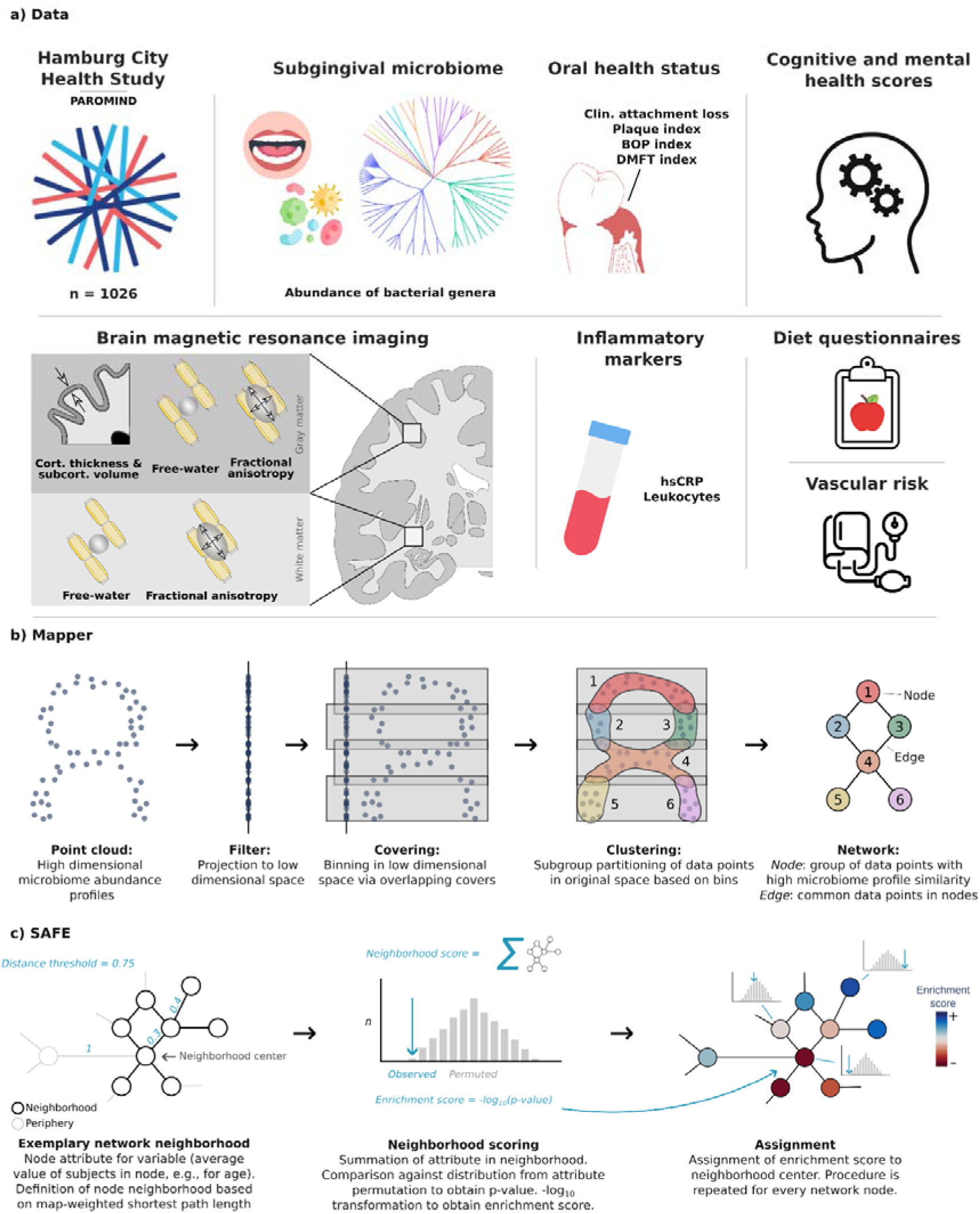
Methodology. a) Population-based data from the Hamburg City Health Study were used including subgingival microbiome abundance, assessments of oral health status, cognitive function and mental health, multimodal brain MRI, circulating inflammatory markers, diet questionnaires, and vascular risk measures. b) We applied the Mapper algorithm to transform the high-dimensional genus-level subgingival microbiome profiles into an interpretable low-dimensional network representation. Mapper consists of multiple steps: (1) Inputting participant-level genus abundance; (2) Projecting data points to a low-dimensional space using a filter function for dimensionality reduction; (3) Dividing the low-dimensional space into overlapping covers, each containing a subset of data points; (4) Clustering data points within each cover based on their distances in the original high-dimensional space; (5) Constructing a network from the clustering results, where each node represents a cluster of participants, and links between nodes indicate shared participants between clusters. Modified from Liao et al.^61^ c) Spatial Analysis of Functional Enrichment (SAFE) was used to annotate the network derived via Mapper, identifying regions significantly enriched with specific attributes (e.g., systolic blood pressure). SAFE involves the following steps: (1) Computing node attributes by averaging variables of interest across participants within each node; (2) Defining the local neighborhood by identifying all nodes within a maximum distance threshold from the center node, with distance measured by map-weighted shortest path length (MSPL); (3) Calculating a neighborhood score by summing the attribute values of neighboring nodes; (4) Computing a p-value by comparing the empirical neighborhood score against a randomly permuted distribution, achieved through random node-to-attribute reassignments while preserving network topology; (5) Assigning an enrichment score to the neighborhood center using -log10 transformation of the multiple testing-corrected p-value. This procedure is repeated for each node, resulting in an enrichment map indicating where attributes are higher or lower than expected by chance. Modified from Baryshnikova et al.^62^ *Abbreviations*: BOP index = bleeding on probing index; clin. = clinical; cort. = cortical; DMFT index = decayed/missing/filled teeth index; hsCRP = high-sensitivity c-reactive protein; subcort. = subcortical.

**Table 1.** Sample characteristics.

| Table 1. Sample characteristics |  |
| --- | --- |
| Metric | Value<sup>a</sup> |
| Age, years | 63.72 ± 8.17 (1,026) [range: 46 - 78] |
| Female, n (% female) | 438 (42.7) |
| Education, ISCED | 2.42 ± 0.58 (999) |
| <u>Oral health measures</u> |  |
| Clinical attachment loss | 2.60 ± 0.83 (842) |
| Plaque index | 19.22 ± 26.30 (830) |
| Bleeding on probing index | 15.38 ± 19.91 (1,011) |
| DMFT index | 19.27 ± 4.90 (842) |
| <u>Cognitive scores</u> |  |
| Animal Naming Test | 24.66 ± 6.67 (977) |
| Mini Mental State Exam | 27.85 ± 1.68 (973) |
| Multiple Choice Vocabulary Intelligence Test | 31.42 ± 3.43 (838) |
| Trail Making Test A, sec | 39.82 ± 14.07 (925) |
| Trail Making Test B, sec | 89.44 ± 37.66 (917) |
| Word List Recall | 7.70 ± 1.88 (949) |
| <u>Mental health scores</u> |  |
| PHQ-9 | 3.21 ± 3.61 (1,025) |
| PHQ-15 | 4.27 ± 3.90 (1,025) |
| Geriatric Depression Scale | 1.83 ± 2.20 (914) |
| GAD-7 | 2.41 ± 2.90 (1,025) |
| <u>Global neuroimaging features</u> |  |
| Free-water (gray matter) | 0.50 ± 0.05 (909) |
| Free-water (white matter) | 0.31 ± 0.03 (909) |
| Tissue fractional anisotropy (gray matter) | 0.19 ± 0.01 (909) |
| Tissue fractional anisotropy (white matter) | 0.41 ± 0.01 (909) |
| Mean cortical thickness, mm | 2.63 ± 0.11 (986) |
| Mean subcortical volume, ml | 3188.31 ± 313.66 (986) |
| <u>Inflammatory markers</u> |  |
| hsCRP | 0.25 ± 0.45 (970) |
| Leukocytes | 6.13 ± 1.75 (1,006) |
| <u>Diet</u> |  |
| MEDAS score | 4.46 ± 1.89 (970) |
| DASH score | 4.46 ± 1.06 (970) |
| MIND score | 6.30 ± 1.78 (970) |
| <u>Vascular risk measurements</u> |  |
| Systolic blood pressure | 142.12 ± 20.06 (997) |
| Diastolic blood pressure | 83.46 ± 10.55 (997) |
| Body mass index | 26.60 ± 4.32 (995) |
| Smoking, % | 173 (16.9) |
| Triglycerides | 117.67 ± 75.68 (1,007) |
| Cholesterol | 208.28 ± 40.83 (1,008) |
| LDL | 121.56 ± 36.53 (997) |
| HDL | 63.51 ± 19.00 (1,008) |
| HbA1c | 5.63 ± 0.54 (1,007) |

Abbreviations: DMFT index = decayed/missing/filled teeth index, HDL = high density lipoprotein, hsCRP = high sensitivity c-reactive peptide, LDL = low density lipoprotein, ISCED = International Standard Classification of Education, mm = millimeters, sec = seconds
<sup>a</sup>Presented as median [IQR] (N)
a) Data

### A similarity network of subgingival bacterial communities

We applied an unsupervised, topology-based technique to abundance data of 85 different bacterial genera to infer a low-dimensional network representation of oral microbiome similarity. The resulting microbiome similarity network consisted of 577 nodes – representing participant groups with highly similar subgingival microbiome compositions – and 10,230 edges – connecting nodes that share at least one participant. The network represents a map of inter-individual variability capturing transitions in microbial composition across the cohort. For a visualization of the network see *figure 2a*.

**Figure 2.**
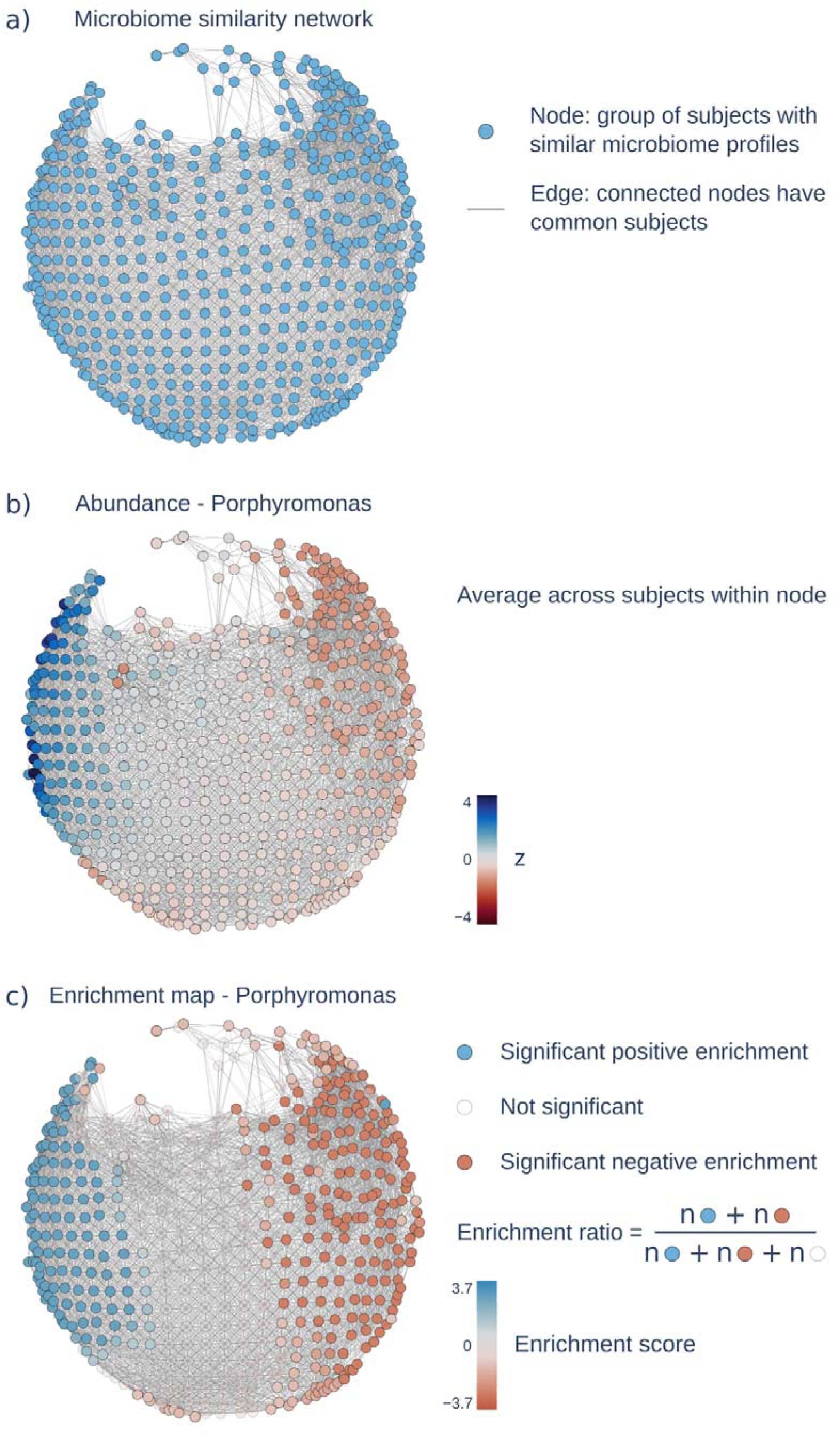
Microbiome similarity network based on genus-level abundance. a) Microbiome similarity network obtained by applying the Mapper algorithm to the abundances of oral microbiome genera consisting of 577 nodes and 10230 edges. Nodes represent groups of participants with similar microbiome profiles. Edges connect nodes that share common participants. Within the network, proximity signifies microbiome profile similarity among participants. b) Exemplary network distribution of z-scored *Porphyromonas* abundance. *Porphyromonas* was selected given its high relevance in periodontal disease.^95^ c) Enrichment map of *Porphyromonas* abundance. Nodes with positive enrichment scores are colored blue indicating a higher *Porphyromonas* abundance of the node and its neighborhood than expected by random permutation. Nodes with negative enrichment scores are colored red indicating lower *Porphyromonas* abundance than expected by permutation. Non-significant nodes are shown in a lighter shade. The enrichment ratio represents a measure of how strongly a phenotype is enriched on a network. It is computed by dividing the count of significantly enriched nodes by all nodes.

In an enrichment analysis, we examined whether the microbiome similarity network captures inter-individual differences in participant traits. Therefore, we computed enrichment scores, which are node-level indices that indicate whether participant traits are significantly higher or lower than expected by chance in specific regions, i.e., participant clusters, of the network. Enrichment scores were obtained for genus-level abundance data as well as all non-microbiome phenotypes. Furthermore, we calculated the enrichment ratio (= number of significantly enriched nodes / total number of nodes) based on the enrichment scores for each phenotype quantifying the overall amount of enrichment on the microbiome similarity network. This measure indicates how strongly the network-based organization of participants reflects inter-individual variance in a respective phenotype. Refer to *figure 2b and 2c* for an exemplary display of *Porphyromonas* abundance and the corresponding enrichment map on the microbiome similarity network.

### Enrichment analysis of genus-level abundance

Microbiome abundance showed significant enrichment across all detected genera ranging from 42.5% to 87.7%. *Figure 3a* displays the enrichment ratios alongside mean relative abundance and phylogenetic associations. We performed a dominance analysis by identifying for each network node the bacterial genus with the highest enrichment scores (*figure 3b*). Of the 85 investigated genera, 15 showed the highest enrichment score for at least 5 nodes. Distribution of dominant genera in the network representation of oral microbiome similarity followed a horizontal pathogenicity gradient: Bacterial genera with strongest enrichments at the *left* end of the microbiome similarity network were periodontitis-associated taxa including *Fusobacterium* (n_nodes_ = 208), *Campylobacter* (65), Treponema (13), *Dialister* (8), *Saccharibacteria* (TM7) [G-5] (6) and *Porphyromonas* (5). In the center of the network, *Aggregatibacter* (18), *Gemella* (8), *Capnocytophaga* (6) and *Leptotrichia* (5) exhibited the highest enrichment scores. At the right end, genera with strongest enrichments were of low periodontal pathogenicity or related to other dental diseases including *Streptococcus* (126), *Veillonella* (42), *Neisseria* (26), *Rothia* (17) and *Haemophilus* (7). For a dominance analysis visualization only considering

**Figure 3.**
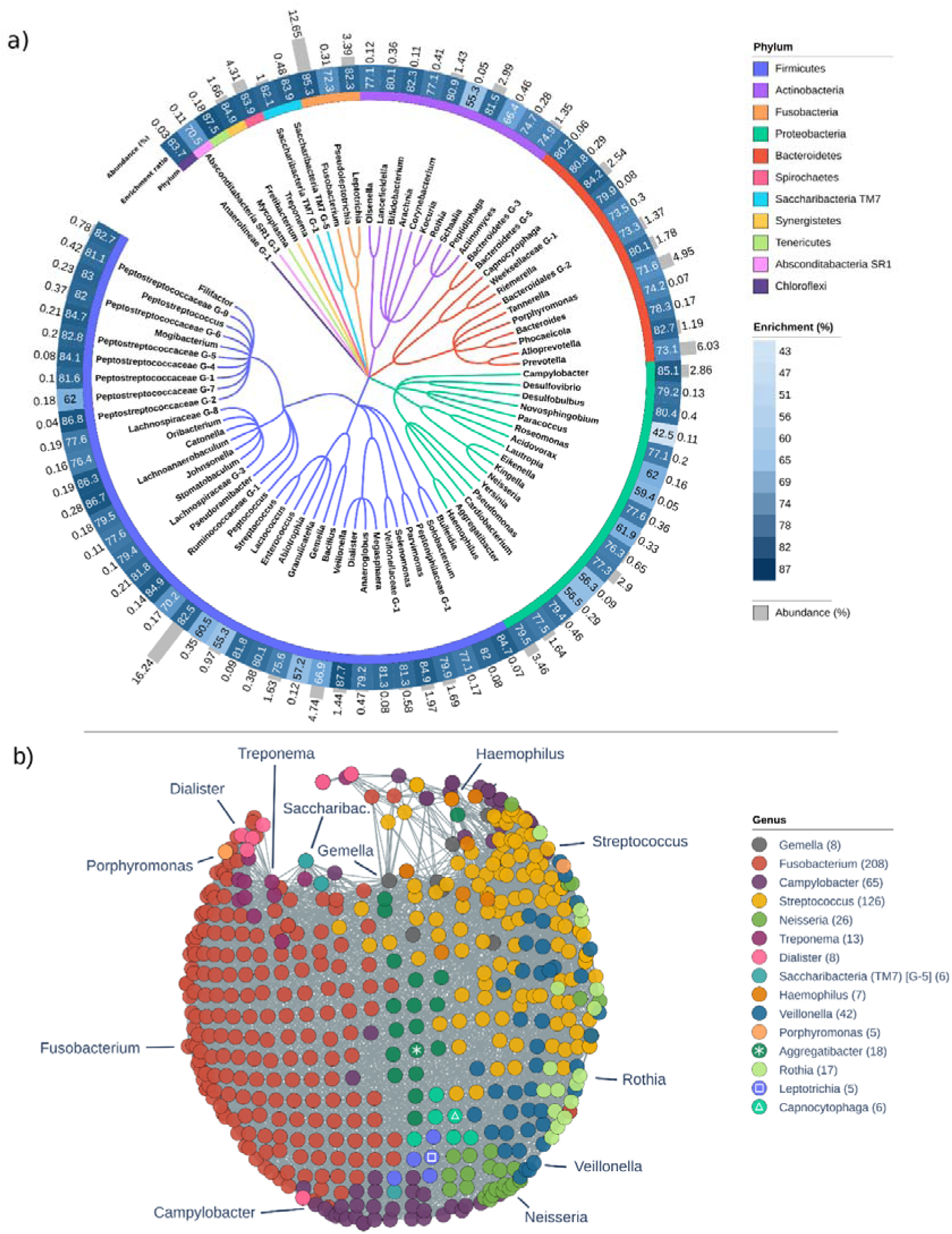
Enrichment analysis of microbiome phenotypes. a) Circular phylogenetic tree at the level of oral microbiome genera. The inner band shows phyla with corresponding coloring of the tree, the mid band displays the enrichment ratio and the outer band shows overall relative abundance. The phylogenetic tree was visualized using iTOL (v6).^96^ b) Dominance analysis: Nodes of the microbiome similarity network are colored to represent the bacterial genus with the highest enrichment score. Genera with dominance in five or more nodes are shown. *Abbreviations*: GM = gray matter, WM = white matter.

### Association of the microbiome configuration with host phenotypes

All non-microbiome phenotypes showed significant enrichment on the microbiome similarity network including measures of oral health status, cognition, mental health, brain structure, circulating inflammatory markers, diet, vascular risk factors and demographics (*figure 4*). The top 10 ranking non-microbiome phenotypes were leukocytes (78.0%), plaque index (77.3%), verbal fluency (75.4%), general cognitive ability (74.7%), smoking behavior (74.5%), Mini Mental State Exam (73.8%), bleeding on probing (BOP) index (72.6%), clinical attachment loss (69.5%), Memory (69.2%) and age (68.6%) (*figure 4a*). A complementary analysis using envfit and adonis confirmed these results: Among the phenotypes with the top 10 highest enrichment ratios, all showed a significant association (p_FDR_ < 0.05) with subgingival microbiome profiles (*supplementary figure S1)*. Moreover, the enrichment ratio was significantly correlated with the adjusted R² resulting from envfit (Spearman ρ = 0.78, p < 0.001) and adonis (Spearman ρ = 0.83, p < 0.001).

**Figure 4.**
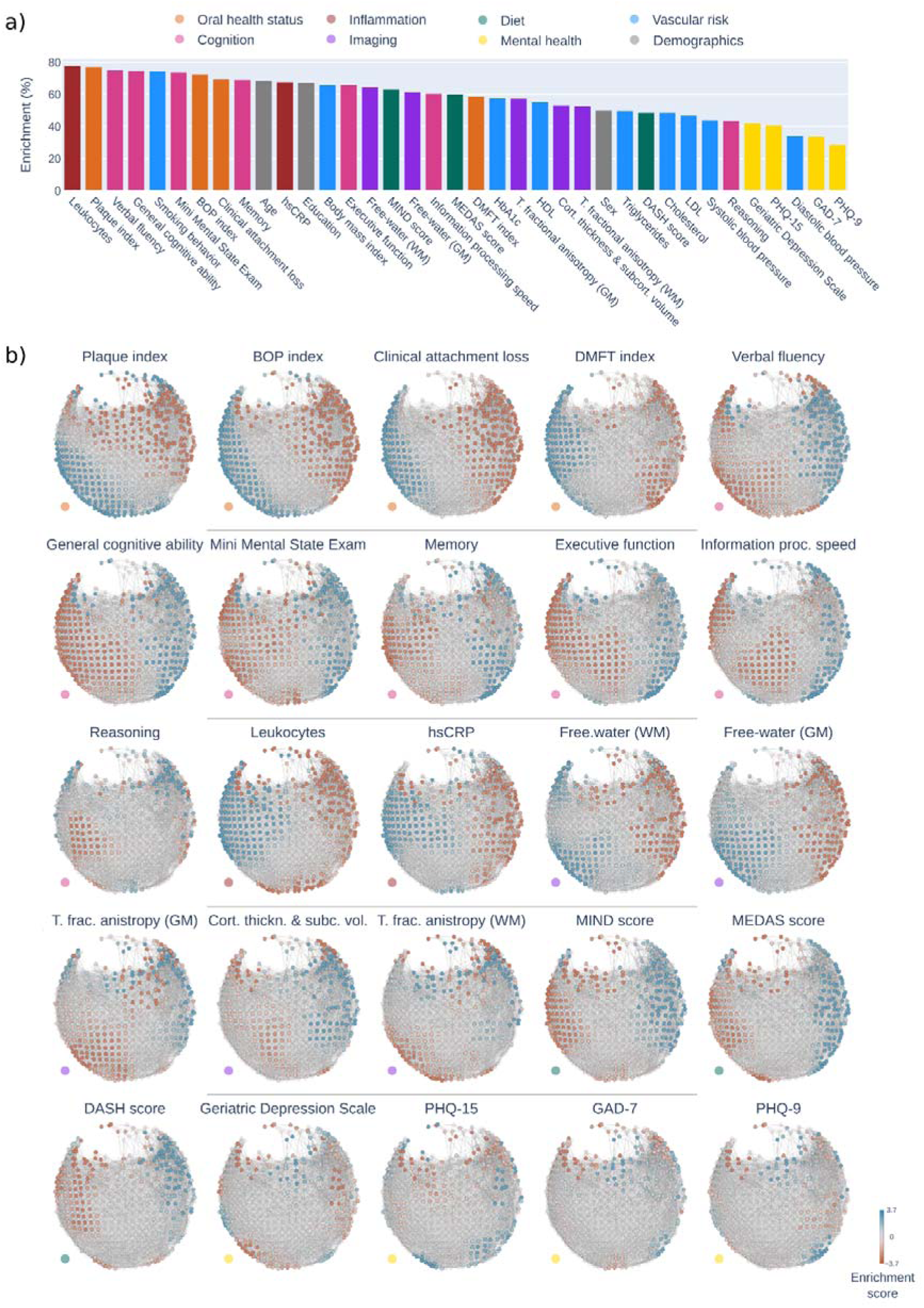
Enrichment analysis of non-microbiome phenotypes. a) The bar plot displays the enrichment ratio (= significantly enriched network nodes / all network nodes) of non-microbiome phenotypes. The enrichment ratio reflects how strongly a phenotype’s variance is captured by the topology of the microbiome network obtained via Mapper. The bars of the plot are colored by variable category. b) Enrichment maps: Enrichment scores (-log_10_-transformed p-values) are mapped on the microbiome similarity network. Nodes with positive enrichment scores, indicating a higher neighborhood score than expected by chance, are colored blue, while nodes with negative enrichment scores, indicating a lower neighborhood score than expected by chance, are colored red. Non-significant nodes are shown in a lighter shade. Non-microbiome phenotypes are first ordered by domain (denoted by colored dots in the lower-left) and then by enrichment ratio. Enrichment maps for demographics, vascular risk factors and genus-level abundances are not displayed and can be found in the online supplement (https://osf.io/vqj8m/)*. Abbreviations*: BOP index = bleeding on probing index; DMFT index = decayed/missing/filled teeth index; GM = gray matter, hsCRP = high sensitivity c-reactive peptide; LDL = low density lipoprotein, T. fractional anisotropy = tissue fractional anisotropy; WM = white matter.

To further disentangle the contributions of non-microbiome variables, we performed a forward model selection (ordiR2step) to identify the set of phenotypes that best explain the variance in microbiome composition (*supplementary figure S2*). Among significantly associated, clinical attachment loss was the strongest contributing factor, explaining 1.64% of the variance, followed by smoking behavior (+0.59%) and the BOP index (+0.54%), gray matter free-water (+0.23%), DMFT index (+0.18%), Plaque index (+0.18%), Leukocytes (+0.15%), Sex (+0.06%), and Mini Mental State Exam (+0.07%). Adding the remaining non-microbiome phenotypes explained an additional 0.04%. Collectively, the final model explained approximately 3.68% of the total variance in the microbiome data.

Based on the enrichment maps, we assessed inter-participant differences of phenotypes. Phenotype enrichment maps are presented in *figure 4b*. Most participant traits varied along the microbiome similarity network in a linear left-right trajectory aligning with the pathogenicity gradient identified via dominance analysis.

Put differently, there was a gradient of enrichment from left to right that reflected the transition of participant characteristics. Participants at the left end of this gradient exhibited higher severity of clinical periodontitis, elevated circulating inflammatory markers, older age, a higher percentage of smokers (indicated by significant positive enrichment), as well as lower cognitive function, lower brain structural integrity, a less healthy diet, a lower percentage of females, and lower education levels (indicated by significant negative enrichment). Conversely, the right end of the gradient featured participants with opposite traits: lower severity of clinical periodontitis, lower circulating inflammatory markers, younger age, fewer smokers, as well as higher cognitive performance, higher brain structural integrity, a healthier diet, a higher percentage of females, and higher education levels. Mental health scores and vascular risk factors beyond smoking displayed a non-linear enrichment pattern not aligned with the pathogenicity gradient.

### Co-enrichment analysis

Principal component analysis of enrichment scores revealed that enrichment patterns of phenotypes differed along two dominant axes of inter-phenotype variation explaining 55.69% (principal component 1, PC1) and 17.49% (principal component 2, PC2) of variance, respectively. Phenotypes with similar enrichment patterns co-localized in the principal component space formed by these axes (*figure 5, supplementary figure S3*). Phenotypes with enrichment patterns indicating increasing values from right end of the network to left were localized on the left extreme of the principal component space including periodontitis-associated bacterial genera, clinical oral health measurements, circulating inflammatory markers, smoking behavior and white matter free-water measured by diffusion-weighted MRI. Phenotypes increasing left to right were located on the right including health-associated bacterial genera, cognitive performance measurements, diet scores, cortical thickness and subcortical volume as measured by MRI. Phenotypes showing a non-linear enrichment pattern or transition pattern in the left-right right-left orientation were localized in the middle including mental health scores and vascular risk measurements apart from smoking. For a heatmap depicting the Spearman correlation of enrichment scores for genus-level abundance and non-microbiome phenotypes see *figure 6*. For the same co-enrichment heatmap indicating genus-genus correlation as well as correlation between non-microbiome phenotypes see *supplementary figures S4 and S5*.

**Figure 5.**
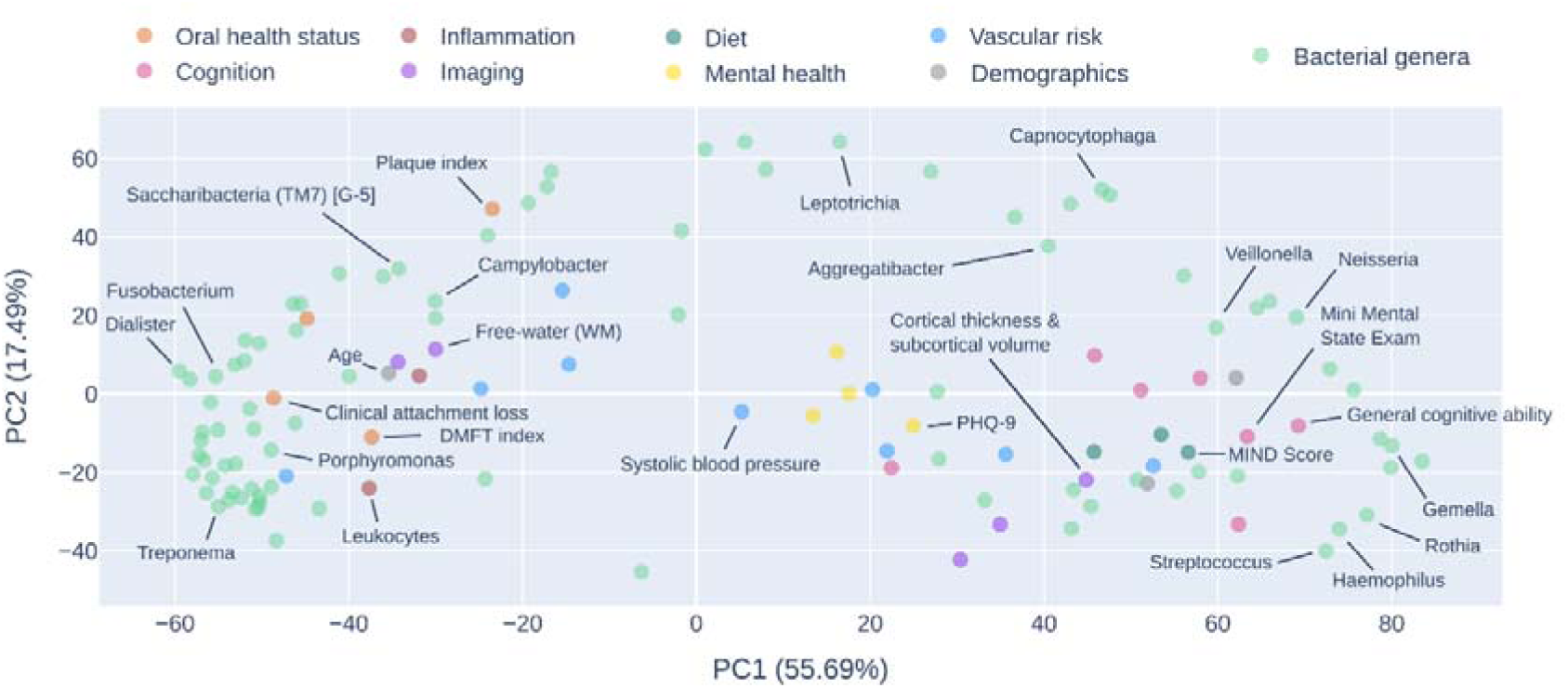
Principal component analysis of enrichment scores. The plot illustrates the phenotypes’ location in principal component space, where proximity suggests similarity in enrichment patterns. Points are color-coded by phenotype category. Dominant genera and selected non-microbiome measures from each phenotype group were highlighted with annotations. For a fully annotated scatter plot, refer to the *supplementary materials*.

**Figure 6.**
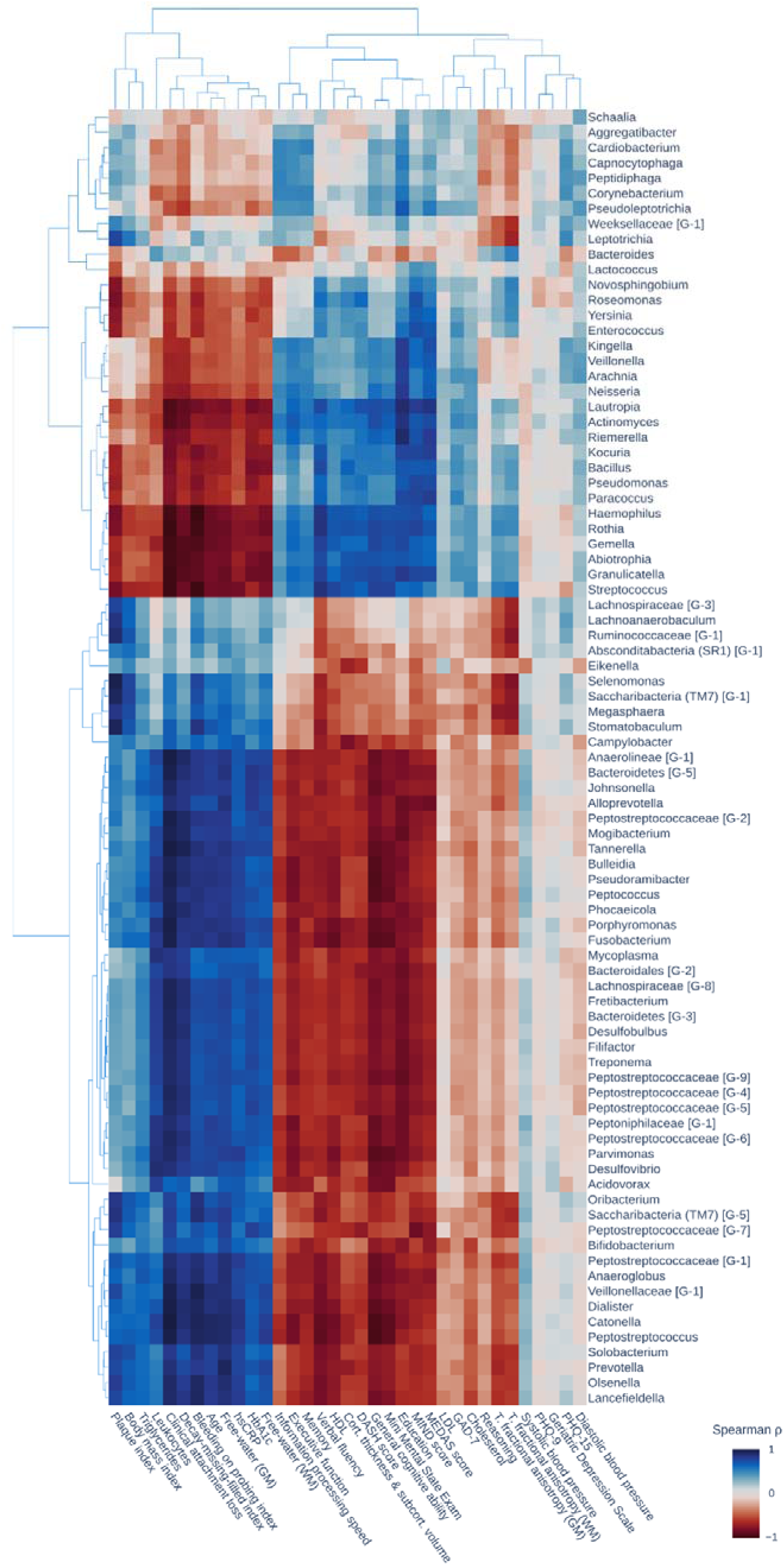
Co-enrichment heatmap. The heatmap presents the Spearman correlation of enrichment scores for genus-level abundance and non-microbiome phenotypes. The order is determined by hierarchical clustering of the enrichment scores. *Abbreviations*: BOP index = bleeding on probing index, DMFT index = decayed/missing/filled teeth index, GM = gray matter, LDL = low density lipoprotein, T. fractional anisotropy = tissue fractional anisotropy, WM = white matter.

### Microbiome similarity groups differ in periodontal disease, inflammatory markers, cognition and diet

Applying k-means clustering on the microbiome similarity network topology resulted in an unsupervised separation of the sample into two participant groups along the pathogenicity gradient. The following statistics to characterize the identified participant groups were adjusted for potential confounders including age, sex, education and vascular risk factors. The procedure is illustrated in *figure 7a*. Participants assigned to nodes in both groups were excluded from the analysis (n_excluded_=137).

**Figure 7.**
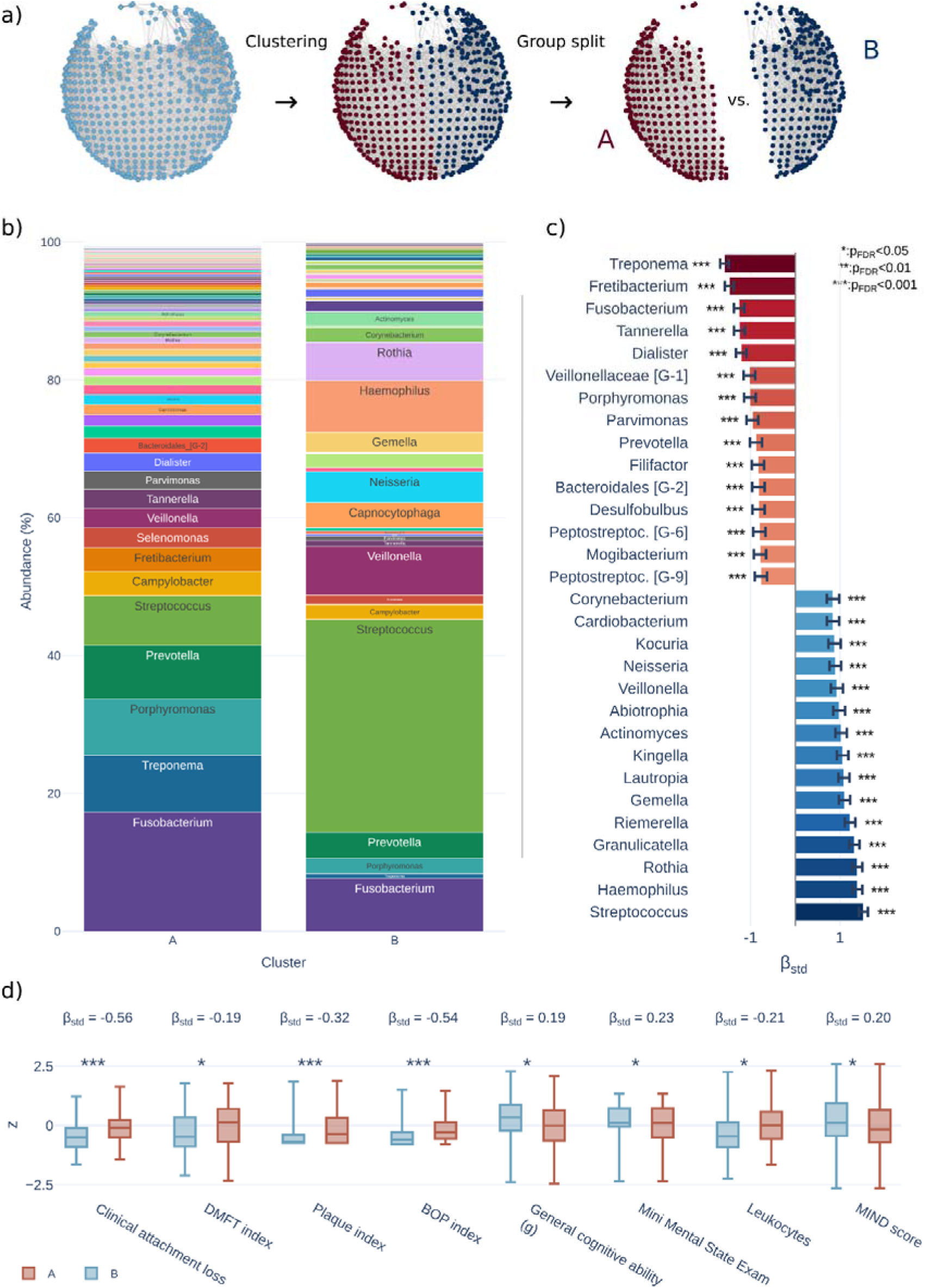
Group analysis. a) Approach: The nodes of the microbiome similarity network were divided into two distinct groups (“A” and “B”) using k-means clustering. Participants were categorized based on these groupings, with individuals present in both groups being excluded from the analysis (n = 137). The two participant groups were then statistically compared using linear regression analysis, adjusting for age, sex, education and vascular risk factors. b) Relative abundance of bacterial genera within groups. c) Top 15 positive and negative differences in microbiome genera: Negative regression coefficients (higher in the “A” group) are shown in red, while positive coefficients (higher in the “B” group) are shown in blue. Significance is indicated by asterisks. For a bar plot displaying coefficients of all investigated genera see *supplementary materials*. d) Shown are box plots for significant associations. Box plot colors correspond to the groups. Statistical significance level is indicated by asterisks. For box plots of the remaining non-microbiome phenotypes see *supplementary materials*. *Abbreviations*: β_std_ = standardized beta coefficient for the group variable. BOP index = bleeding on probing index, DMFT index = decayed/missing/filled teeth index, GM = gray matter; *Peptostreptoc.* = *Peptostreptococcaceae*; WM = white matter.

We compared genus-level abundance to identify bacterial genera that showed significant differences between groups (*figure 7b* and *7c*). Of the 85 tested genera, 8 showed no significant group differences (*supplementary figure S6*). Top 15 bacterial genera which were significantly higher in the “A” group were *Treponema* (β_std_ = - 1.58), *Fretibacterium* (β_std_ = -1.47), *Fusobacterium* (β_std_ = -1.25), *Tannerella* (β_std_ = - 1.25), *Dialister* (β_std_ = -1.20), *Veillonellaceae* [G-1] (β_std_ = -1.03), *Porphyromonas* (β_std_ = -1.01), *Parvimonas* (β_std_ = -0.96), *Prevotella* [G-2] (β_std_ = -0.87), *Filifactor* (β_std_ = - 0.83), *Bacteroidales* [G-2] (β_std_ = -0.82), *Desulfobulbus* (β_std_ = -0.82), *Peptostreptoccocaceae* [G-6] (β_std_ = -0.80), *Mogibacterium* (β_std_ = -0.79), and *Peptostreptoccocaceae* [G-9] (β_std_ = -0.77) (all p_FDR_ < 0.001). Top 15 bacterial genera which were significantly higher in the “B” group were *Streptococcus* (β_std_ = 1.52), *Haemophilus* (β_std_ = 1.39), *Rothia* (β_std_ = 1.38), *Granulicatella* (β_std_ = 1.32), *Riemerella* (β_std_ = 1.23), *Gemella* (β_std_ = 1.10), *Lautropia* (β_std_ = 1.09), *Kingella* (β_std_ = 1.06), *Actinomyces* (β_std_ = 1.03), *Abiotrophia* (β_std_ = 0.98), *Veillonella* (β_std_ = 0.93), *Neisseria* (β_std_ = 0.90), *Kocuria* (β_std_ = 0.88), *Cardiobacterium* (β_std_ = 0.84), and *Corynebacterium* (β_std_ = 0.84) (all p_FDR_ < 0.001).

In addition, the “A“ group showed significantly higher clinical severity of periodontitis, higher circulating inflammatory markers, lower cognitive performance and lower brain structural integrity compared to the “B” group (*figure 7d*): a significantly higher clinical attachment loss (β_std_ = -0.56, p_FDR_ < 0.001), DMFT index (β_std_ = -0.15, p_FDR_ = 0.047), plaque index (β_std_ = -0.32, p_FDR_ < 0.001), bleeding on probing index (β_std_ = - 0.53, p_FDR_ < 0.001), leukocytes (β_std_ = -0.21, p_FDR_ = 0.014) as well as a significantly lower general cognitive ability (β_std_ = 0.18, p_FDR_ = 0.026), Mini Mental State Exam score (β_std_ = 0.22, p_FDR_ = 0.014) as well as MIND score (β_std_ = 0.20, p_FDR_ = 0.026). The remaining cognitive scores, diet scores and hsCRP showed no significant differences (*supplementary figure S7*). Moreover, brain structural indices and mental health scores also showed no significant differences. These results were stable across models reflecting different sequential adjustment steps (*supplementary figure S8*).

### Sensitivity analysis

To assess the robustness of our result we performed two-fold sensitivity analyses.

First, we conducted repeated enrichment analyses on randomly selected subsamples, reducing the sample size from 100% to 10% in 1% decrements, with 100 random samples at each step (*supplementary figure S9*). Throughout this process, the Spearman correlation of enrichment ratios (covering both non-microbiome and microbiome phenotypes) demonstrated high robustness (Spearman ρ > 0.8) to different sample compositions until the sample size was reduced to approximately 80%. Additionally, the Adjusted Rand Index (ARI) remained high (ARI > 0.8), indicating strong consistency in microbiome similarity network-based group assignments from k-means clustering, until the sample size was reduced to approximately 70%.

Second, to evaluate the potential influence of pipeline design choices on our findings, we reanalyzed the data using 17 different pipeline configurations, varying the components and parameters of the topological data analysis (*supplementary table S10*). The results demonstrated high robustness, with Spearman correlations between enrichment ratios from the original and alternative configurations averaging 0.82 ± 0.03. Furthermore, the ARI for group assignments derived from k-means clustering via the original pipeline, compared to those from different configurations, remained consistently high at 0.84 ± 0.06.

## Discussion

The oral cavity hosts the second most diverse microbiome in the human body, following the gut.^22^ Interactions within these oral microbial communities, along with host factors, affect both oral and systemic health.^23^ In this study, we present a comprehensive population-level analysis examining the association between the subgingival microbiome and multi-domain brain health-related phenotypes ranging from cognitive functions, mental health, brain structure, inflammatory blood biomarkers, to dietary behavior and vascular risk factors. Using 16S rRNA sequencing and topological data analysis, we inferred a microbiome similarity network that revealed a continuous pathogenicity gradient, mapping individuals based on periodontal microbiome profiles. Leveraging this network, we identified associations of periodontitis-related microbial communities with multiple brain health phenotypes: Individuals with higher abundance of periodontitis-related taxa exhibited significantly lower cognitive performance, lower MIND diet scores as well as increased leukocyte counts adjusting for demographics and cardiovascular risk. Our findings highlight several key insights into the oral microbiome-brain axis, potential pathophysiological pathways and clinical implications.

### Topological data analysis reveals a latent axis of periodontitis-related microbial composition

We performed a dominance analysis to determine which bacterial genera are particularly abundant within specific participant clusters of the microbiome similarity network. Based on this analysis, we show that the microbiome similarity network captured interindividual variations and a gradient of microbial compositions from periodontitis-associated taxa to health-associated taxa (*Figure 3b*): Periodontitis-associated genera such as *Porphyromonas*, *Fusobacterium*, *Treponema, Saccharibacteria (TM7)* and *Campylobacter* were enriched at the left part of the network, consistent with their established roles in biofilm formation, immune modulation, and periodontal tissue destruction.^9,24–27^ Also positioned at this pathogenic end were bacteria of the genus *Dialister*, which have only recently been associated with periodontitis and could be a key periodontal pathogen, warranting further research into its mechanisms.^28^ In contrast, genera such as *Streptococcus*, *Haemophilus*, *Rothia, Veillonella and Neisseria* were enriched at the right part, reflecting their roles in maintaining an oral health equilibrium, their overall low periodontal pathogenicity or their association with oral conditions different from periodontitis, such as dental caries.^29–31^ Enrichment ratios for bacterial genera were overall high, affirming that the observed interindividual differences were relevantly captured by the microbiome similarity network.

These findings indicate that periodontitis emerged as the most parsimonious explanation for the topology of the microbiome similarity network, with participants harboring higher abundances of periodontitis-related taxa clustering at one end, and those with higher abundances of other genera clustering at the other. Put differently, the identified network revealed a latent axis and continuum of periodontitis (referred to as pathogenicity gradient) mirroring the pathogenicity spectrum of periodontitis and suggesting that the disease constitutes a key driver of the observed interindividual variance of microbiome compositions. This evidence demonstrates that a topological data analysis-based approach can uncover subtle yet biologically meaningful patterns of disease severity in large-scale, highly complex datasets.

### Microbial compositions are linked to non-microbiome phenotypes

The microbiome similarity network offers a means to integrate complex, high-dimensional microbiome data with comprehensive health information by mapping non-microbial phenotypes on the network and statistically testing their association with the network structure. We observed that the investigated non-microbiome phenotypes differentially enriched on the network, implying varying degrees of association with microbial configuration. These enrichment patterns ranged from linear enrichments correlated or anti-correlated with the pathogenicity gradient (e.g., leukocytes) – where variance in these variables coincided with changes in the abundance of periodontitis-related taxa – to non-linear patterns indicating weaker or more complex associations with the pathogenicity gradient (e.g., PHQ-9) (*Figure 4b*). Co-enrichment analyses further detailed these observations, showing that microbial and non-microbial phenotypes with visually overlapping enrichment patterns occupied similar positions in principal component space and exhibited strong correlations in their enrichment scores (*Figures 5* and *6; supplementary figures S3-S5*).

To quantitatively compare these different analytical approaches, we tested for and discovered a strong and significant correlation between the SAFE enrichment ratio (the primary outcome of our topological analysis) and the adjusted R² from both adonis (Spearman’s ρ=0.83, p<0.001) and envfit (Spearman’s ρ=0.78, p<0.001). This finding demonstrates that the patterns discovered through our novel topological framework are quantitatively consistent with the global effect sizes identified by traditional methods. Crucially, while the overall trend is consistent, the specific rankings of variables differed between methods (*supplementary figure S1*). We argue that this is not a contradiction but rather an informative result that highlights the unique sensitivities of each approach. The topological analysis can detect nuanced or non-linear patterns (e.g., as demonstrated for the mental health scores) that are not fully captured by a single global variance statistic. The traditional approaches facilitate contextualizing our results with previous analyses.

Among the phenotypes most closely aligned with the pathogenicity gradient were the plaque index, clinical attachment loss, and bleeding on probing index, reinforcing periodontitis as the primary pathology captured by the network.^32,33^ A forward model selection analysis confirmed that clinical indicators of periodontitis ranked among the strongest contributors to the variance in microbiome composition (*supplementary figure S2*). These findings highlight not only the complex covariance structure of subgingival microbiome composition but also the potential of genus-level abundance data as a promising biomarker source, reflecting a broad range of clinical and lifestyle factors.

This analysis also highlighted significant, though smaller, contributions from brain structure (gray matter free-water), cognitive performance (Mini Mental State Exam), and systemic inflammation (leukocytes). This suggests, while oral health status is the primary factor associated with microbial profiles, these profiles also reflect systemic and brain-related health measures. Jointly, host phenotypes explained 3.68% of variance in microbiome composition. This level of explained variance, while low, is consistent with findings from other major population studies and confirms that these host factors have a measurable impact on the microbial community.^34,35^

### Periodontal dysbiosis and brain health

The mechanisms behind the relationship between periodontitis and brain health are multifaceted and remain to be fully elucidated. Our analysis sought to address these complexities by integrating data on multiple brain health phenotypes into a single analysis framework.

Notably, multiple markers of cognitive performance (verbal fluency, general cognitive ability, Mini Mental State Exam, Memory) and systemic inflammation (leukocyte counts, hsCRP) were among non-oral phenotypes with strongest enrichment ratios (*Figure 4a*) and varied in tandem with the pathogenicity gradient from one network end to the other (*Figure 4b*). Specifically, participants with higher abundance of periodontitis-associated taxa and poorer oral health exhibited significantly lower cognitive performance and elevated systemic inflammation after adjusting for covariates (*Figure 7c* and *7d*). Confirming these topological findings, our forward model selection analysis also revealed significant associations between the microbiome and brain structure (gray matter free-water), cognitive performance (Mini Mental State Exam), and systemic inflammation (leukocytes) (*supplementary figure S2*). Importantly, these results build on prior reports linking shifts in oral microbiota to cognitive changes among dementia patients and demonstrate that such associations also manifest in a healthy population of non-demented, middle-aged individuals.^18,36^ Furthermore, our findings align with earlier analyses indicating relationships between clinical markers of periodontitis and cognitive health.^37–40^

Our analysis highlights associations between brain health and the abundance of various bacterial genera (*Figure 7b* and *7c*). These findings are consistent with prior research implicating genera such as *Porphyromonas*, *Treponema*, *Fusobacterium*, and *Prevotella* in pathways potentially relevant to Alzheimer’s disease. Prominently, experimental evidence from animal models demonstrated that *Porphyromonas gingivalis* contributes to Alzheimer’s pathology by contributing to the formation of amyloid-β, neurofibrillary tangles and neuroinflammation.^11,13^ Furthermore, postmortem studies report a higher abundance of *Porphyromonas gingivalis* in the brain samples of AD patients compared to non-demented controls, with intracerebral presence of the bacterium being related to a six-fold increased risk of AD.^11,41^ *Treponema denticola* has been demonstrated to induce tau hyperphosphorylation and neuroinflammatory processes in mice.^42^ The serum levels of antibodies against *Prevotella intermedia* and *Fusobacterium nucleatum* are significantly increased in Alzheimer’s disease patients.^15^ Additionally, elevated *Prevotella intermedia* abundance is associated with *APOE4*-carrier status, and *Fusobacterium* species have been linked to cerebrovascular lesions indicative of small vessel pathology.^17,43^ Our findings align with these results, indicating that the prevalence of these pathogenic bacteria in the subgingival biofilm corresponds to reduced brain health in cognitively normal individuals. At the same time, we found genera not yet reported in relation to brain health – *Fretibacterium*, *Tannerella*, *Dialister*, *Parvimonas, Filifactor, Peptostreptococcaceae,* and *Mogibacterium* – warranting further experimental research to determine whether they independently drive cognitive deficits or merely coincide with other pathogenic species.

Our findings emphasize the role of periodontitis-related taxa in systemic inflammation and possible connections to neurodegenerative pathways. We observed that individuals positioned at the pathogenic end of the oral microbiome similarity network exhibited significantly higher leukocyte levels after covariate adjustment (*Figure 7d*), indicating presence of systemic inflammation. This aligns with prior studies linking clinically diagnosed periodontitis to elevated systemic inflammatory markers.^44^ Pathomechanistically, such responses can stem from dense leukocytic infiltration during gingival inflammation, the stimulation of bone marrow for sustained inflammatory cell production, or the systemic response to dissemination of pathogens from ulcerated gingival tissues.^44^ Importantly, we present novel evidence linking periodontitis-associated microbial compositions to systemic inflammation. Notably, this systemic inflammation was accompanied by reduced cognitive performance, supporting the hypothesis that chronic oral infections contribute to both systemic inflammation and cognitive disease. Although our findings cannot prove causality due to the cross-sectional nature of the study design, they align with the inflammatory hypothesis of Alzheimer’s disease, suggesting that immune responses triggered by oral pathogens may disrupt the blood-brain barrier and activate microglia, thereby driving neurodegeneration.^45^

Previous analyses also highlight an association between periodontal health and brain structure.^20,46,47^ However, the global brain structural measures we assessed did not show significant group differences after adjustment for covariates. We speculate that this could be because any potential structural effects related to the oral microbiota in this cohort are subtle or regionally specific, rather than global, and thus not captured by the overall brain metrics used. Future analysis should therefore focus on assessing localized brain structural changes, potentially using voxel-based or region-of-interest methods, to determine if subtle or regionally specific associations exist with oral microbial profiles.

In contrast to cognitive measures, mental health scores displayed weaker associations with the pathogenicity gradient, suggesting more nuanced or less direct links with periodontitis-related microbiome composition. These associations did not persist after adjusting for demographic and cardiovascular risk factors. We interpret these results as pointing to a more specific link of periodontitis to cognitive rather than mental health. Nevertheless, prior studies have reported connections between anxiety, depression, and clinical periodontitis or oral microbiome composition.^48^ Longitudinal research will be essential to elucidate how these factors collectively influence oral and brain health.

### Influence of diet and vascular risk factors on oral and brain health

We investigated the role of nutritional behavior as a key factor influencing oral microbiome composition.^49^ Enrichment analysis revealed that adherence to cognitively beneficial diets, such as the MEDAS, MIND, and DASH diets,^50–52^ aligned with the pathogenicity gradient: participants with greater abundance of periodontitis-related taxa tended to adhere less to these diets. Notably, after adjusting for covariates, individuals with a higher abundance of periodontitis-related genera exhibited significantly lower adherence to the MIND diet. These findings corroborate previous results from the HCHS, which demonstrated that higher adherence to DASH, MEDAS and an anti-inflammatory dietary was associated with lower odds of periodontal disease.^53^ Our findings suggest that dietary patterns known to promote cognitive health may also shape the oral microbiome toward a less pathogenic composition. Given that the MIND diet specifically emphasizes foods with anti-inflammatory and neuroprotective properties,^54^ it is plausible that such nutritional patterns could influence both oral and brain health through microbial community shifts and immune modulation. A healthier diet may foster a healthier oral microbiome based on its emphasis on fiber over simple sugars selectively promoting beneficial microbes while limiting resources for harmful ones.^55^ Furthermore, it could reduce gingival inflammation through anti-inflammatory compounds, making the environment less hospitable to pathogens potentially disrupting brain health.^56^ Longitudinal and interventional studies are needed to disentangle these pathways and clarify causal relationships between diet, oral microbial ecology, and brain health.

Among demographic and vascular risk factors, smoking behavior was the only factor clearly associated with the pathogenicity gradient. Smoking has been documented to promote the colonization of periodontal pathogens, facilitate biofilm formation, and compromise the immune response, thereby exacerbating periodontal disease and systemic inflammation.^57^ Our findings highlight the role of smoking on promoting dysbiosis and inflammation and positions it as a direct contributor to the pathogenicity gradient and brain health phenotypes.

### Clinical implications

Our findings not only shed light on the intricate systemic pathophysiology of periodontal dysbiosis and brain health, but they also hint at potential avenues of clinical utilization. Our study reveals an association between subgingival microbiome signatures and variance in cognitive performance and brain structure that could enhance early screening measures for risk of cognitive decline and improve targeted recruitment of individuals at critical early stages of cognitive impairment and dementia. Particularly, identifying microbial community changes before emergence of cognitive symptoms could enhance diagnostics by providing microbiome-based markers as a convenient complement to existing dementia biomarkers. Regarding future therapeutic interventions, microbiome signatures of early periodontal dysbiosis may guide the development of oral microbiome-directed therapies to slow or prevent dementia progression.^58^ However, the definitive role of microbiome biomarkers in cognitive disorders is yet to be determined and large-scale longitudinal as well as interventional studies are required for moving in this direction.

### Strengths and limitations

Strengths of this work lie in its considerable sample size; comprehensive taxonomic profiling of the oral microbiome; in-depth phenotyping of clinical and lifestyle data; as well as a novel, robust data analysis pipeline that unifies multiple complex data domains into a single analysis framework. Specifically, the large, population-based cohort provided the microbial diversity to model a continuous disease gradient, while its deep phenotyping enabled our topological approach to map complex microbial-phenotype associations.

However, our study also exhibits limitations. First, an important limitation of our study is its cross-sectional design, which precludes definitive causal inference and the assessment of temporal relationships. The relationship between oral and brain health is increasingly understood not as a simple unidirectional link, but as a complex, bidirectional relationship, with evidence supporting causality in both directions. The hypothesis of reverse causality posits a valid and mechanistically plausible pathway where individuals with poorer cognitive function might face challenges in maintaining optimal oral hygiene, which could in turn influence their periodontal health status and subgingival microbiome composition. Previous work systematically reviewing longitudinal studies acknowledges that observed associations between poor periodontal health and cognitive decline are at least partly due to this effect.^59^ Conversely, the forward pathway is supported by a growing body of experimental research. This external evidence demonstrates that periodontal pathogens can trigger neuroinflammation and contribute to neurodegenerative pathology, lending significant weight to a potential contributing causal role for oral dysbiosis in affecting brain health.^11,13^ Future longitudinal follow-up of the PAROMIND cohort is planned and will be essential to explicitly disentangle the temporal dynamics of this complex oral-brain relationship and to track the progression from subclinical microbial and brain changes to overt clinical outcomes. Second, data on participant relatedness or household sharing were unavailable; these factors are known to influence microbiome similarity and could therefore potentially act as unmeasured confounders in our analyses. Third, our analysis relied on pooled subgingival samples from the deepest periodontal pocket in each quadrant. While this standardized approach was designed to capture a representative profile of sites with active disease, it may obscure potentially important site-specific differences in microbial composition across the oral cavity.

Finally, our reliance on 16S rRNA gene sequencing limits the current analysis primarily to taxonomic composition, precluding direct assessment of microbial functional potential or metabolic activity. While computational tools can predict function from 16S data, integrating such inferred analyses was beyond the scope of this initial broad, multi-domain study focused on taxonomic associations. Our planned next steps include exploring computationally inferred functions (e.g., using PICRUSt2^60^) on the existing dataset. However, for direct functional characterization, investigation of specific metabolic pathways, resolution of strain-specific taxonomic variations (including virulence factors), and identification of potential non-bacterial microbiome members, future studies utilizing high-resolution methods such as shotgun metagenomics will be essential to gain deeper mechanistic insights into the oral microbiome-brain axis.

## Conclusion

Drawing on a comprehensive investigation integrating multi-domain data with cutting-edge analysis techniques we characterized latent associations of periodontitis-related subgingival microbial composition with cognitive function, mental health, brain structural integrity, systemic inflammation, diet and vascular risk in predominantly healthy individuals. Notably, after adjusting for potential confounders, periodontitis-related oral dysbiosis was associated with lower cognitive performance, lower MIND diet adherence, and higher leukocyte counts. As this research field progresses, oral microbiome profiling could contribute to improved dementia risk stratification and guide preventive interventions.

## Materials and methods

### Study design

Our methodology is illustrated in *figure 1*. This analysis integrates different data domains related to oral and brain health into a unified analysis framework: subgingival microbiome composition, oral health status, cognitive function, mental health status, brain structure, circulating inflammatory markers, dietary patterns, vascular risk and demographics. In brief, we employed topological data analysis, specifically the Mapper algorithm, to create a low-dimensional network representation of subgingival microbiome abundance data.^61^ This network positions individuals based on similarities in their subgingival microbiome composition. Subsequently, we conducted an enrichment analysis using Spatial Analysis of Functional Enrichment (SAFE).^62^ This analysis statistically assesses regions of the network derived via Mapper to identify where specific phenotypes are significantly higher or lower than expected by chance. This approach allowed us to examine regional differences in phenotypes, revealing whether the network representation captures variance in specific participant traits related to oral and brain health. Finally, we performed a post-hoc group comparison between participants of two distinct participant clusters within the microbiome network, enabling us to assess the link of subgingival microbiome composition and clinical and lifestyle phenotypes while adjusting for relevant covariates.

### Study population

PAROMIND is a cross-sectional study nested within the Hamburg City Health Study (HCHS). The HCHS is a prospective, single-center, population-based cohort study investigating adults aged 45-75 to enhance the detection of major chronic disease risks through extensive clinical and imaging phenotyping.^21^ Participants were included in PAROMIND for periodontal examination and subgingival sampling if they reported no antibiotic use within the preceding three months, had no requirement for endocarditis prophylaxis, and possessed more than two remaining teeth. The study protocol includes assessments of oral health (including collection of gingival crevicular fluids for microbiome analyses), cognition, mental health, diet, vascular risk, brain MRI, and blood sampling. For each participant, the clinical assessments and biological sampling were generally completed during a single comprehensive baseline visit; the separate brain MRI appointment followed shortly thereafter, ensuring reasonable contemporaneity between these measures for cross-sectional analysis.

### Ethics statement

PAROMIND and the HCHS were approved by the local ethics committee of the Landesärztekammer Hamburg (State of Hamburg Chamber of Medical Practitioners, PV5131). The conduct of PAROMIND is governed by ethical guidelines of Good Clinical Practice (GCP), Good Epidemiological Practice (GEP) and the Declaration of Helsinki.^63^ Written informed consent was obtained from all participants investigated in this work.

### Molecular analysis and phenotyping of the oral microbiome

Oral microbiome phenotyping followed a standardized procedure targeting the subgingival environment within periodontal pockets. This specific environment was chosen because it contains the subgingival biofilms most directly implicated in periodontitis pathogenesis, distinguishing its microbial community from those in other oral sites like saliva or supragingival plaque. During the dental examination, gingival crevicular fluid samples were collected from periodontal pockets with sterile paper points. Two samples per participant were obtained: (1) from a single deep periodontal pocket and (2) one pooled sample (4 paper points) from the deepest periodontal pockets per quadrant. Each paper point remained in situ for 15 seconds. Subsequently, samples were placed in a sterile 2ml-Eppendorf tube and stored at - 80°C in the HCHS biobank. For the following molecular analysis, only pooled samples were further processed.

Sequencing was performed at the Institute of Clinical and Molecular Biology in Kiel, Germany. All samples were processed with negative and positive controls. The amount of DNA per sample was quantified prior to amplification to ensure it met the required minimum of 50 ng, which is particularly important for low-biomass samples like gingival crevicular fluid. Bacterial composition was determined via 16S rRNA Illumina sequencing. DNA Isolation was performed using the DNA extraction kit from Innuprep Analytics (Analytik Jena AG, Überlingen, Germany). For the initial lysis with lysozyme and mutanolysin (3 mg lysozyme, 100 U mutanolysin, in 200 µl Tris EDTA buffer), all samples were incubated at 37°C for 10 min and further isolation was performed according to the manual with elution volumes of 100 µl. For 16S rDNA sequencing of isolated DNA, variable regions V3 and V4 of the 16S rRNA gene were amplified using the primer pair 341F (5‘-CCTACGGGAGGCAGCAG-3‘) and 806R (5‘-GGACTACHVGGGTWTCTAAT-3’) 27F-338R in a dual barcoding approach.^64,65^ Agarose gel electrophoresis was used to verify resulting PCR products before normalization using the SequalPrep Normalization Plate Kit (Thermo Fischer Scientific, Waltham, MA, USA), pooling and sequencing on the Illumina MiSeq with v3 2×300bp chemistry (Illumina Inc., San Diego, CA, USA). Demultiplexing after sequencing was based on 0 mismatches in the barcode sequences.

Data processing was performed using the DADA2 workflow for big datasets (v. 1.10.42, https://benjjneb.github.io/dada2/bigdata.html), resulting in abundance tables of amplicon sequence variants (ASVs). For this, all sequencing runs were handled separately and finally collected in a single abundance table per dataset, which underwent chimera filtering. ASVs underwent taxonomic annotation using the Bayesian classifier provided in DADA2 and using the expanded Human Oral Microbiome Project (eHOMD) version V15.23. Samples with less than 10,000 sequences were not considered for further analysis. Downstream analyses were performed at the genus-level because this taxonomic rank provided a suitable balance, enhancing statistical power across the large cohort, improving interpretability and comparability with existing literature, and reducing data dimensionality while still providing sufficient resolution for the study’s objectives. Genera that were observed with a frequency of less than 0.1% of all genera detected in a sample were discarded.

### Oral health assessment

A certified study nurse assessed probing depth and gingival recession at six sites of the tooth (mesio-buccal, buccal, disto-buccal, disto-palatinal, palatinal and mesio-palatinal). The clinical attachment loss (CAL) was calculated (CAL = probing depth + gingival recession).^66^ The BOP index was determined by probing 2 sites per tooth (vestibular, oral) and expressed as a percentage of bleeding sites. The number of bleeding sites on probing is a reliable and consistent measure to assess the degree of gingival inflammation. Measurements were completed with a standard periodontal probe (PCP 15, Hu-Friedy, Chicago, IL, USA). Additionally, the DMFT index was calculated. The oral health assessment followed a standardized protocol for the reporting of the prevalence and severity of periodontal diseases.^67^

### Cognitive and mental health assessments

Cognitive testing was conducted using the extended version of the Consortium to Establish a Registry for Alzheimer’s Disease Neuropsychological Assessment Battery (CERAD-NP/Plus).^68^ A trained study nurse administered all tests. For this analysis, we considered cognitive scores measuring executive function (Trail Making Test B), information processing speed (Trail Making Test A), memory (Word List Recall Test), reasoning (Multiple Choice Vocabulary Intelligence Test B), verbal fluency (Animal Naming Test), and the Mini Mental State Exam. To ensure higher scores indicated better cognitive performance across all tests, we inverted the results of Trail Making Test A and B. Subsequently, we performed a principal component analysis (PCA) on all individual test scores. Following previous procedures, the first principal component, which accounted for the greatest variance (40.5%), was defined as a measure of general cognitive ability (g) (for details see *supplementary figure S11*).^69^ According to the principal component loadings, higher values of this measure corresponded to lower cognitive performance; thus, it was also inverted. Furthermore, participants underwent mental health assessments via established questionnaires for depression (PHQ-9, Geriatric Depression Scale), somatic symptom severity (PHQ-15), and anxiety (GAD-7).^70–72^

### Neuroimaging of brain macro- and microstructure

Neuroimaging markers representing different aspects of macro- and microstructural brain integrity were computed based on T1-weighted and diffusion-weighted MRI following previous procedures.^73^ After cortical surface reconstruction and subcortical segmentation based on T1-weighted images with FreeSurfer (v. 6.0.1), cortical thickness and subcortical volume were estimated representing morphometric measures of neurodegenerative processes.^74,75^ The mean cortical thickness and mean subcortical volume were z-scored. Subsequently, the two resulting z scores were averaged to obtain a single summary measure reflecting cortical thickness and subcortical volume. Following preprocessing of the diffusion-weighted images, free-water imaging was employed to compute free-water which quantifies the amount of extracellular water as well as tissue fractional anisotropy reflecting neurite architecture and integrity.^76,77^ The free-water and tissue fractional anisotropy values were then averaged across cortical and subcortical gray matter voxels, as well as white matter voxels, to obtain global measures of gray and white matter, respectively. For a detailed account on the acquisition protocol, quality assessment, preprocessing and computation procedures on the different imaging measures see *supplementary text S12*.

### Diet

The dietary behavior of all participants was assessed using a validated food frequency questionnaire with 102 items, developed for the European Prospective Investigation into Cancer and Nutrition Study (EPIC).^78^ The adherence to different dietary patterns was measured based on the food frequency questionnaire scores, including the Mediterranean diet (MEDAS diet), the Dietary Approaches to Stop Hypertension (DASH diet), and the Mediterranean-DASH Intervention for Neurodegenerative Delay (MIND diet).^50–52^ Adherence to the Mediterranean diet was determined using the German version of the Mediterranean Diet Adherence Screener, which assigns a score of 0 or 1 to 14 specific food items, producing a total adherence score ranging from 0 (no adherence) to 14 (maximum adherence).^50^ The DASH diet was evaluated using a previously established scoring method that assigns a score of 0, 0.5, or 1 to each of 10 items, resulting in an adherence score between 0 (no adherence) and 10 (maximum adherence).^52^ Finally, adherence to the MIND diet was calculated according to standard procedures: scores of 0, 0.5, or 1 to are assigned 10 healthy and 5 unhealthy food items, culminating in a total adherence score ranging from 0 (no adherence) to 15 (maximum adherence).^51^

### Topological data analysis

We implemented a topological data analysis pipeline that integrates two key components: (1) the Mapper algorithm, which performs an unsupervised reconstruction of a topological network based on genus-level abundance data, capturing microbiome composition similarity, and (2) SAFE, which conducts statistical tests to examine the relationship between the network’s structure and different phenotypes.^61,62^ This method effectively combines dimensionality reduction with topological insights, offering a powerful tool for understanding the intrinsic geometry of high-dimensional microbiome data and its relationships with other data domains. The analysis was performed in *python* v3.8.1 based on the packages *NetworkX* v2.2 (https://github.com/networkx/networkx), *safepy* (https://github.com/baryshnikova-lab/safepy), *scikit-learn* v1.5.1 (https://github.com/scikit-learn/scikit-learn) and *tmap* v1.2 (https://github.com/GPZ-Bioinfo/tmap) as well as *R* v4.4.0 based on the package *vegan v2.6-6.1*.^61,62^ Data visualization was based on *plotly* v5.22 (https://github.com/plotly/plotly.py) and iTOL v6 (https://itol.embl.de/). HTML versions of many presented plots can be found on OSF allowing interactive data exploration (https://osf.io/vqj8m/).

### Mapper: Reconstruction of the microbiome similarity network

Genus-level subgingival microbiome abundance data served as input to the Mapper algorithm, a topological data analysis technique that simplifies complex high-dimensional data by constructing a topological network capturing essential relationships and patterns in the data. This network preserves the data’s underlying topological and geometric structure by positioning participants with similar subgingival microbiome profiles nearby. Conceptually, this representation is analogous to a topographical map that reveals the essential features of a landscape. Importantly, the network can represent non-linear associations that conventional linear techniques might miss. Mapper has previously been used to analyze the dynamic organization of brain function,^79,80^ the shape of genetic data in breast-cancer patients,^81^ biomolecular folding pathways,^82^ brain structure in patients with fragile X syndrome,^83^ and neuronal data from the visual cortex.^84^

The applied Mapper pipeline comprises multiple analysis steps to reconstruct the topological network (for an illustration see *figure 1b*): filtering, covering, clustering and network reconstruction.^85^ First, as the filtering step, we conducted a principal coordinate analysis of the Aitchison distance of genus-level abundance (n_participants_ x n_genera_).^86^ To account for the compositional nature of the data and handle zero values, the distance was calculated using the robust Aitchison method.^87^ By that, we obtained two components – also called lenses – capturing the major axes of variation in the microbial community composition across the participants. Based on these axes, overlapping covers were defined (overlap = 1.5, resolution = 30) to segment the data into overlapping bins, each representing a local region of inter-individual variation. Unsupervised clustering of datapoints within each bin was performed using Hierarchical Density-Based Spatial Clustering of Applications with Noise (HDBSCAN, epsilon threshold = 0.95).^88^ By this, nodes are obtained that represent participant groups with similar configuration of the subgingival microbiome. Participants can belong to multiple nodes with the number varying per participant. Lastly, the network reconstruction was accomplished by connecting clusters sharing common participants. Not all datapoints are retained by Mapper resulting in the omission of some participants (n_not retained_ = 109).

### SAFE: Enrichment analysis

SAFE is an annotation technique for biological networks that enables the computation and statistical assessment of local enrichment for specific phenotypes.^62^ In our work, we performed SAFE on the microbiome similarity network derived using the Mapper algorithm to identify regions within the network that are significantly enriched for specific participant traits. Specifically, we investigated the enrichment of genus-level microbiome abundance, oral health measures (clinical attachment loss, plaque index, bleeding on probing index, DMFT index), cognitive scores (general cognitive ability, Animal Naming Test, Mini Mental State Exam, Multiple Choice Vocabulary Intelligence Test B, Trail Making Tests A and B, Word List Recall), mental health scores (PHQ-9, PHQ-15, Geriatric Depression Scale, GAD-7), imaging measures (cortical thickness and subcortical volume, gray matter free-water, white matter free-water, gray matter tissue fractional anisotropy, white matter tissue fractional anisotropy), circulating inflammatory markers (hsCRP, leukocytes), diet scores (MEDAS score, DASH score, MIND score), vascular risk factors (systolic and diastolic blood pressure, body mass index, smoking behavior, blood triglycerides, cholesterol, low density lipoprotein, high density lipoprotein, HbA1c), and demographics (age, sex, education).

Following previous analyses leveraging SAFE,^62,89^ the microbiome similarity network was spring-embedded, i.e., nodes in the network were positioned so that those with connections are placed closer together, while repulsive forces push non-connected nodes apart, resulting in a visually balanced and interpretable representation of the network’s structure.^90^ Next, we computed node attributes for all phenotypes by averaging the values of each variable across all participants within a node. Subsequently, enrichment scores were calculated for each node following a four-step process (*figure 1c*). First, we defined the local neighborhood by identifying all nodes within a maximum distance threshold of 0.75 from the central node. The distance was measured using the map-weighted shortest path length (MSPL).^62^ Second, we calculated a neighborhood score by summing the attribute values of neighboring nodes. Third, we computed a p-value by comparing the empirical neighborhood score against a distribution derived from 5000 permutations. Permutations were performed by randomly reassigning attributes to nodes while preserving the network topology.^62^ The resulting p-value was corrected for multiple comparisons across all phenotypes. Finally, we assigned an enrichment score to the neighborhood center by applying a -log10 transformation to the corrected p-value. Given the 5000 permutations, the maximal enrichment score is -log10(1/5000) = 3.70, with -log10(0.05) = 1.30 indicating significance. Positive enrichment scores indicate that observed values are higher than the permuted distribution, negative enrichment scores indicate that they are lower than the permuted distribution. This procedure is repeated for each node of the network, resulting in an enrichment map indicating where attributes are higher or lower than expected by chance.

To understand the primary taxonomic patterns shaping the network topology, we performed a dominance analysis alongside examining individual phenotype enrichments. This involved labeling each network node by the single bacterial genus with the highest positive enrichment score. Visualizing this node-level dominance helps to reveal major taxonomic transitions across the network. While highlighting the most strongly enriched genus in each region provides valuable pointers to key drivers, this is a simplification; each node still represents a complex microbial community, not just the dominant genus identified.

### Statistical assessment of microbiome-host associations

To measure how strongly the microbiome similarity network reflects a specific phenotype we computed the enrichment ratio as the number of significantly enriched nodes (corrected p < 0.05) divided by the total number of nodes. A higher enrichment ratio indicates that more nodes are significantly enriched, suggesting that the network’s topology captures a greater extent of a phenotype’s variance.

While the enrichment ratio quantifies the scope of a phenotype’s association across the network’s topology, it is not a direct measure of effect size. To provide a complementary, conventional assessment of effect size, we also tested the association between non-microbiome phenotypes and the overall microbiome configuration using two statistical methods from the vegan package: 1) envfit (n_permutations_ = 5000) testing the linear association between each phenotype and the principal coordinate analysis (PCoA) ordination (all components) of the robust Aitchison distance matrix; 2) permutational multivariate analysis of variance (PERMANOVA, adonis, n_permutations_ = 5000) testing the proportion of variance in the robust Aitchison distance matrix explained by each phenotype individually. All resulting p-values were false discovery rate-corrected. This complementary analysis was only performed for non-microbiome phenotypes and not for the abundance of individual genera.

To quantify the relative contribution of non-microbiome phenotypes while accounting for their overlapping effects, we performed forward model selection using distance-based redundancy analysis (db-RDA, capscale). The analysis was conducted with the ordiR2step function (n_permutations_ = 5000) from *vegan*, using the robust Aitchison distance matrix of the microbiome data as the response variable. Because ordiR2step requires a complete dataset, missing values in the non-microbiome data were imputed using k-nearest neighbors (KNN, n_neighbors_ = 5) prior to model fitting. This imputation was based only on other non-microbiome phenotypes to prevent information leakage from the microbiome data. The procedure sequentially added explanatory variables, retaining only those that significantly improved the model’s fit (p < 0.05 based on 5000 permutations). The adjusted R2 value for each step was used to quantify the variance explained by each selected variable.

To determine whether specific phenotypes co-enrich, i.e., exhibit similar enrichment patterns, we performed an ordination of the enrichment scores using principal component analysis retaining the first two principal components and assessed the pairwise Spearman correlation between the enrichment scores.

### Group analysis

To further examine the relationship between the microbiome similarity network and clinical as well as lifestyle phenotypes, we performed a post-hoc group analysis which allowed us to adjust for relevant covariates. Therefore, nodes of the microbiome similarity network were clustered in two non-overlapping groups using k-Means clustering of the node positions. We chose k-Means as it is widely used and arguably represents the simplest unsupervised clustering technique.^91^ k-Means requires to predefine the number of clusters to assign datapoints to. Given that enrichment analysis indicated that most participant traits varied along the microbiome similarity network in a linear left-right trajectory, i.e., participants on the left end differed from those on the right end, the number clusters was predefined as n_clusters_ = 2 post-hoc.

After the clustering, participants were categorized based on the resulting groupings. Importantly, individuals present in both groups – due to being assigned to nodes in both groups – were excluded from the analysis (n = 137). Given that not all datapoints are retained by the Mapper algorithm during the network reconstruction step, participants that were not represented in the microbiome similarity network were not considered for this analysis. Before the group comparison, a center log-ratio (CLR) transformation was applied to genus-level abundance data. The groups were statistically compared for the phenotypes using multiple linear regression and age, sex and education and vascular risk factors (systolic and diastolic blood pressure, body mass index, smoking behavior, triglycerides, cholesterol, LDL, HDL, HbA1c) were included as covariates:

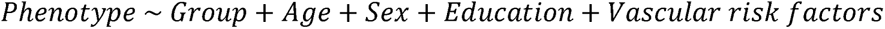

Covariates were selected a priori based on literature demonstrating their potential influence on both oral health/microbiome and brain health indices.^47,92–94^ In addition to this fully adjusted model, we performed an exploratory analysis by building models that sequentially added each covariate, allowing us to map the specific influence of each confounder on the primary associations.

### Sensitivity analyses

To verify the robustness of our results against variations in the sample as well as the analysis pipeline, we conducted a comprehensive sensitivity analysis.

This involved repeating the entire analysis across random subsamples of different sizes. Specifically, we varied the sample sizes from 100% to 10% of the total dataset, in 1% decrements. For each decrement, we randomly sampled subsets 100 times, resulting in a total of 9,000 iterations (90 different sample sizes × 100 random samples per steps). For each iteration, we assessed the robustness of our findings by comparing the results from the subsamples to the original results. The stability of the enrichment analysis results was evaluated by calculating the Spearman correlation of enrichment ratios for the non-microbiome phenotypes and genus-level abundance. In addition to this, we measured the agreement of the participant-group assignments resulting from k-Means clustering used for group analysis employing the Adjusted Rand Index (ARI) which ranges from 0 (no agreement) to 1 (full agreement).

The parameters and components of our analysis pipeline were selected based on established practices, including strategies to optimize sample coverage as described in the tmap documentation, or utilized default software settings. Recognizing that pipeline parameters can influence resulting network characteristics (e.g., node count, connectivity), we conducted a sensitivity analysis using alternative configurations to verify that our results were not biased by these initial design choices. We systematically explored variations in parameter values and pipeline components, altering the Mapper cover overlap from the original 1.5 to 1, 1.2, 1.4, 1.6, 1.8 and 2; the Mapper cover resolution from 30 to 20, 25, 35, 40 and 45; and the Mapper epsilon threshold from 0.95 to 0.99, and 0.90. Additionally, we adjusted the SAFE distance threshold from 0.75 to 0.5 and 0.99, and the SAFE neighborhood radius from 0.1 to 0.05 and 0.15. We adjusted one parameter from the original pipeline per iteration, resulting in a total of 15 iterations. The robustness of our results was evaluated by comparing the findings from the original configuration to those from alternative setups. Matching the approach for the previous sensitivity analysis, we calculated the Spearman correlation of enrichment ratios for clinical and genus abundance phenotypes and the ARI for group assignments from k-Means clustering to assess stability.

## Data Availability

HCHS data can be obtained by qualified researchers on reasonable request to the studys steering committee. The analysis code for this work is publicly available on GitHub (https://github.com/csi-hamburg/oral_microbiome_brain_health). Interactive versions of the plots can be found on OSF (https://osf.io/9ykv5).

## Data availability

Sequencing data generated during this study have been deposited as FASTQ files in the European Nucleotide Archive (ENA) under accession number PRJEB89258. Corresponding non-microbiome phenotype data from the HCHS are not publicly available due to data protection policies that ensure participant confidentiality. However, this data is available to qualified researchers upon reasonable request to the HCHS steering committee. The analysis code for this work is publicly available on GitHub (https://github.com/csi-hamburg/oral_microbiome_brain_health). Interactive versions of the plots can be found on OSF (https://osf.io/vqj8m/).

## Acknowledgments

The authors would like to acknowledge all participants, all cooperation partners, patrons and the Deanery from the University Medical Center Hamburg-Eppendorf for supporting the HCHS. Special thanks are due to the staff at the Population Health Research Department for conducting the study. The publication of its results has been approved by the Steering Board of the HCHS. We also thank the staff of the microbiome and sequencing laboratories of the Institute of Clinical Molecular Biology Kiel for their excellent support.

## Author Contributions

Each author has made a significant contribution to the manuscript and all authors read and approved its final version. We describe contributions to the paper using the CRediT contributor role taxonomy. M.P.: Conceptualization, Data curation, Formal analysis, Investigation, Methodology, Project administration, Resources, Software, Visualization, Writing—original draft, Writing—review & editing; C.W.: Conceptualization, Data curation, Investigation, Methodology, Resources, Writing— original draft, Writing—review & editing; K.B.: Data curation, Methodology, Investigation, Resources, Software, Writing—review & editing; G.H.: Resources, Writing—review & editing; T.B.: Resources, Writing—review & editing; M.A.: Resources, Writing—review & editing; C.M.: Resources, Writing—review & editing; F.L.N.: Data curation, Resources, Writing—review & editing; B.Z.: Resources, Writing—review & editing; J.F.: Resources, Writing—review & editing; J.G.: Resources, Writing—review & editing; S.K.: Resources, Writing—review & editing; R.T.: Resources, Writing—review & editing; C.B.: Methodology, Writing—review & editing; G.T.: Supervision, Funding, Writing—review & editing; B.C.: Conceptualization, Funding, Project administration, Resources, Supervision, Writing—original draft, Writing—review & editing; G.A.: Conceptualization, Funding, Project administration, Resources, Supervision, Writing—original draft, Writing— review & editing.

