## Supplementary materials for "Oral microbiome profiles relate periodontal disease and brain health - the PAROMIND Study"

# -

### Supporting information

Marvin Petersen[1*], Carolin Walther[2*], Katrin Borof[2], Guido Heydecke[3], Thomas Beikler[2], Malik Alawi[4], Christian Müller[4], Felix L. Nägele[1], Birgit-Christiane Zyriax[5], Jens Fiehler[6], Jürgen Gallinat[7], Simone Kühn[7,8], Raphael Twerenbold[9,10,11,12], Corinna Bang[13], Götz Thomalla[1], Bastian Cheng[1#], Ghazal Aarabi[2#]

*[1]Department of Neurology, University Medical Centre Hamburg-Eppendorf, Hamburg, Germany.*

*[2]Department of Periodontics, Preventive and Restorative Dentistry, University Medical Center Hamburg-Eppendorf, Hamburg, Germany*

*[3]Department of Prosthetic Dentistry, University Medical Center Hamburg-Eppendorf, Hamburg, Germany*

*[4]Bioinformatics Core, University Medical Center Hamburg-Eppendorf, Hamburg, Germany*

*[5]Midwifery Science-Health Services Research and Prevention, Institute for Health Services Research in Dermatology and Nursing (IVDP), University Medical Center Hamburg-Eppendorf, Hamburg, Germany*

*[6]Deparment of Neuroradiology, University Medical Center Hamburg-Eppendorf, Hamburg, Germany*

*[7]Department of Psychiatry and Psychotherapy, University Medical Center Hamburg-Eppendorf, Hamburg, Germany*

*[8]Lise Meitner Group for Environmental Neuroscience, Max Planck Institute for Human Development, Berlin, Germany*

*[9]Department of Cardiology, University Heart and Vascular Center, Hamburg, Germany*

*[10]Epidemiological Study Center, University Medical Center Hamburg-Eppendorf, Hamburg, Germany*

*[11]German Center for Cardiovascular Research (DZHK), partner site Hamburg/Kiel/Luebeck, Hamburg, Germany*

*[12]University Center of Cardiovascular Science, University Heart and Vascular Center, Hamburg, Germany*

*[13]Institute of Clinical Molecular Biology, Kiel University, Kiel, Germany*

**shared first authorship*

*[#]shared last authorship*

#### Figure S1 – Results from envfit and adonis

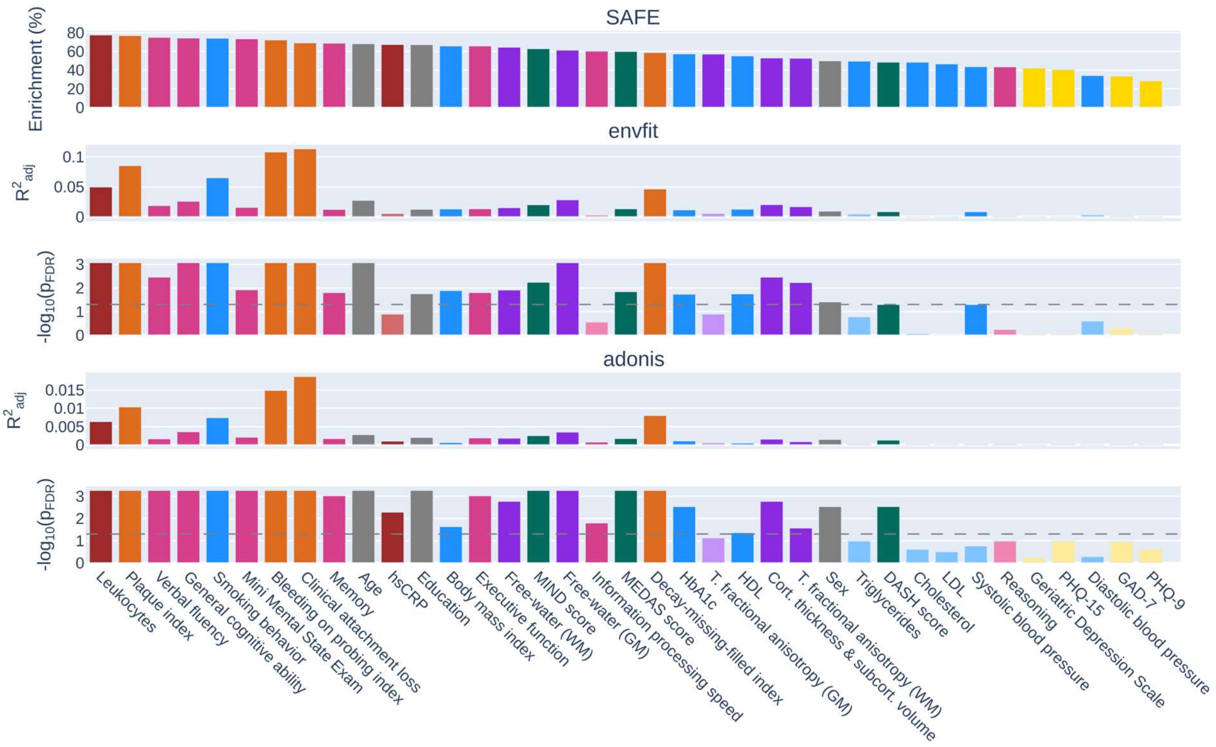

The top bar plot displays the SAFE enrichment ratio (% enrichment) for non-microbiome phenotypes, reflecting how strongly each phenotype's variance is captured by the topology of the microbiome similarity network (obtained using Aitchison distance PCoA as input), as shown in the main manuscript. As complementary approaches, we assessed the association between the same phenotypes and the overall microbiome configuration (genus-level Aitchison distance matrix) using two complementary statistical methods: 1) envfit, testing the linear association between each phenotype and the PCoA ordination (all components) of the Aitchison distance matrix; 2) adonis (PERMANOVA), testing the proportion of variance in the Aitchison distance matrix explained by each phenotype individually. For both methods, we show the effect size, quantified as the adjusted R², and the corresponding statistical significance, displayed as -log10($p_{FDR}$). For all significance plots, the horizontal dashed line indicates the significance threshold of $p_{FDR} = 0.05$. The bars in all plots are colored by variable category.

Summary: Phenotypes exhibiting the highest enrichment ratios (top plot) generally show significant associations across the complementary linear tests (envfit, adonis), confirming their relationship with the overall subgingival microbiome configuration. However, perfect concordance is not observed, potentially reflecting differences between the methods in their distinct statistical approaches and sensitivities to different types of patterns. The topological analysis is designed to detect the specific "shape" of an association and can identify complex, non-linear patterns, which our study found for certain phenotypes (e.g., mental health scores). In contrast, adonis performs a global test of association, and envfit is most sensitive to linear trends. This difference in what each method tests (e.g., local patterns vs. global variance) could explain the lack of perfect concordance.

*Abbreviations*: BOP index = bleeding on probing index; db-RDA = distance-based redundancy analysis; DMFT index = decayed/missing/filled teeth index; GM = gray matter, hsCRP = high sensitivity c-reactive peptide; LDL = low density lipoprotein, PERMANOVA = Permutational multivariate analysis of variance, $R^2_{adj}$ = Adjusted R²; T. fractional anisotropy = tissue fractional anisotropy; WM = white matter.

#### Figure S2 – Forward model selection analysis (ordiR2step)

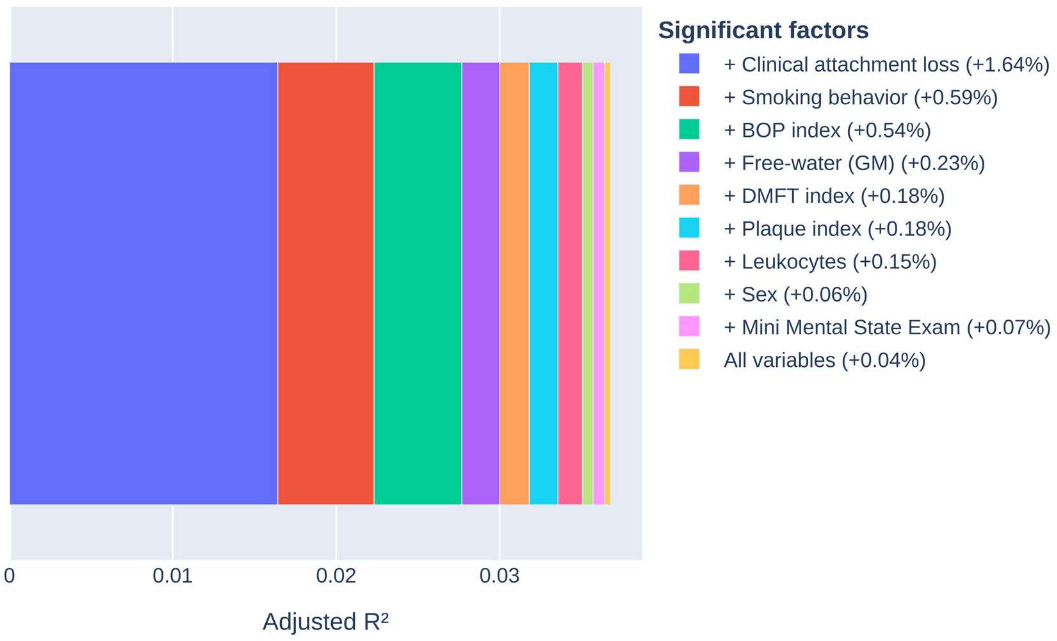

The stacked bar plot displays the cumulative variance (adjusted $R^2$) in the subgingival microbiome explained by significant non-microbiome phenotypes, as determined by forward model selection (ordiR2step).

### Figure S3 – Principal component analysis of network enrichment scores with annotations

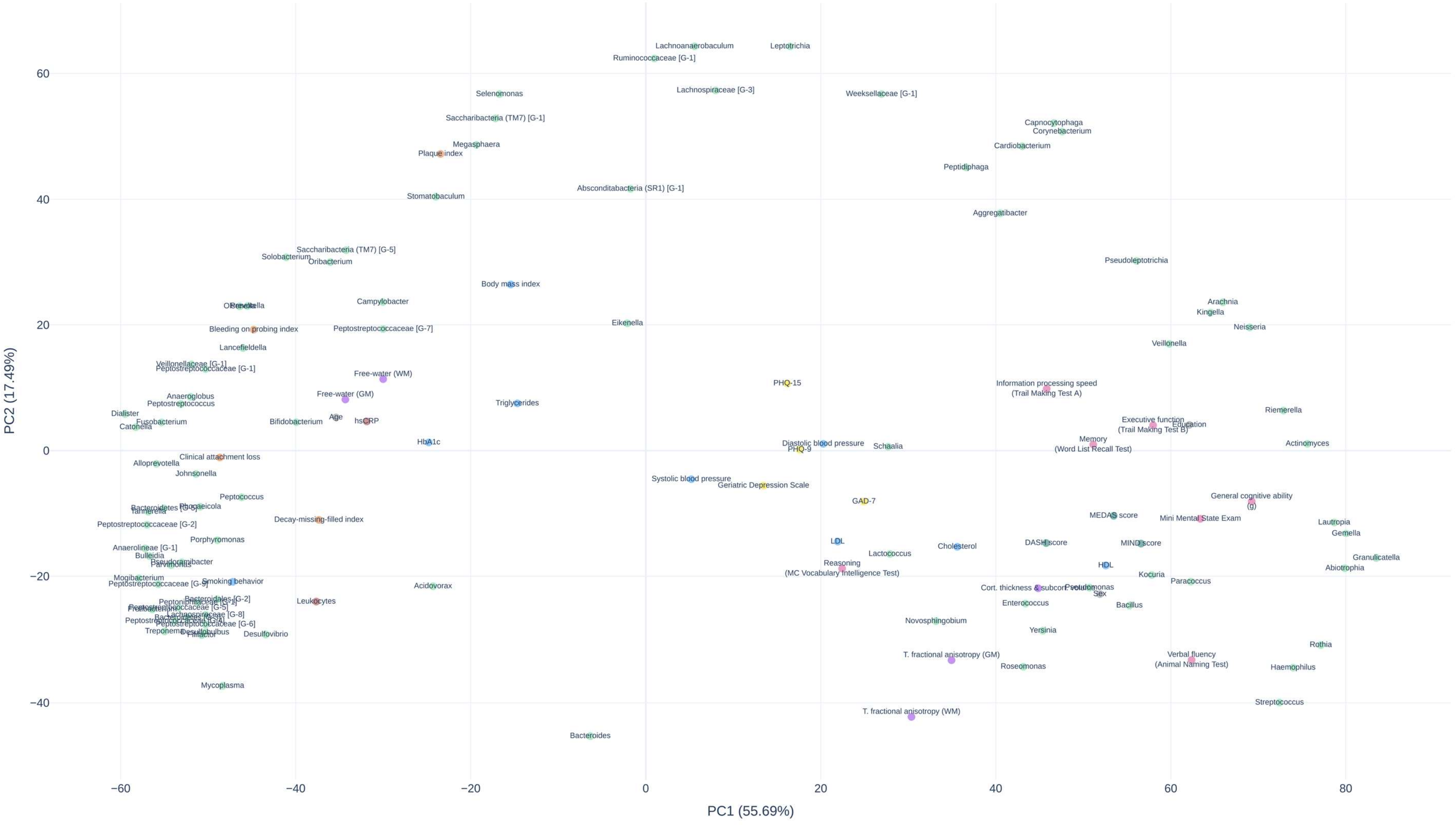

Principal component analysis of enrichment scores. The plot displays phenotypes in principal component space, where proximity indicates similar enrichment patterns. Points are color-coded by phenotype category.

#### Figure S4 – Co-enrichment heatmap of genus-level abundance

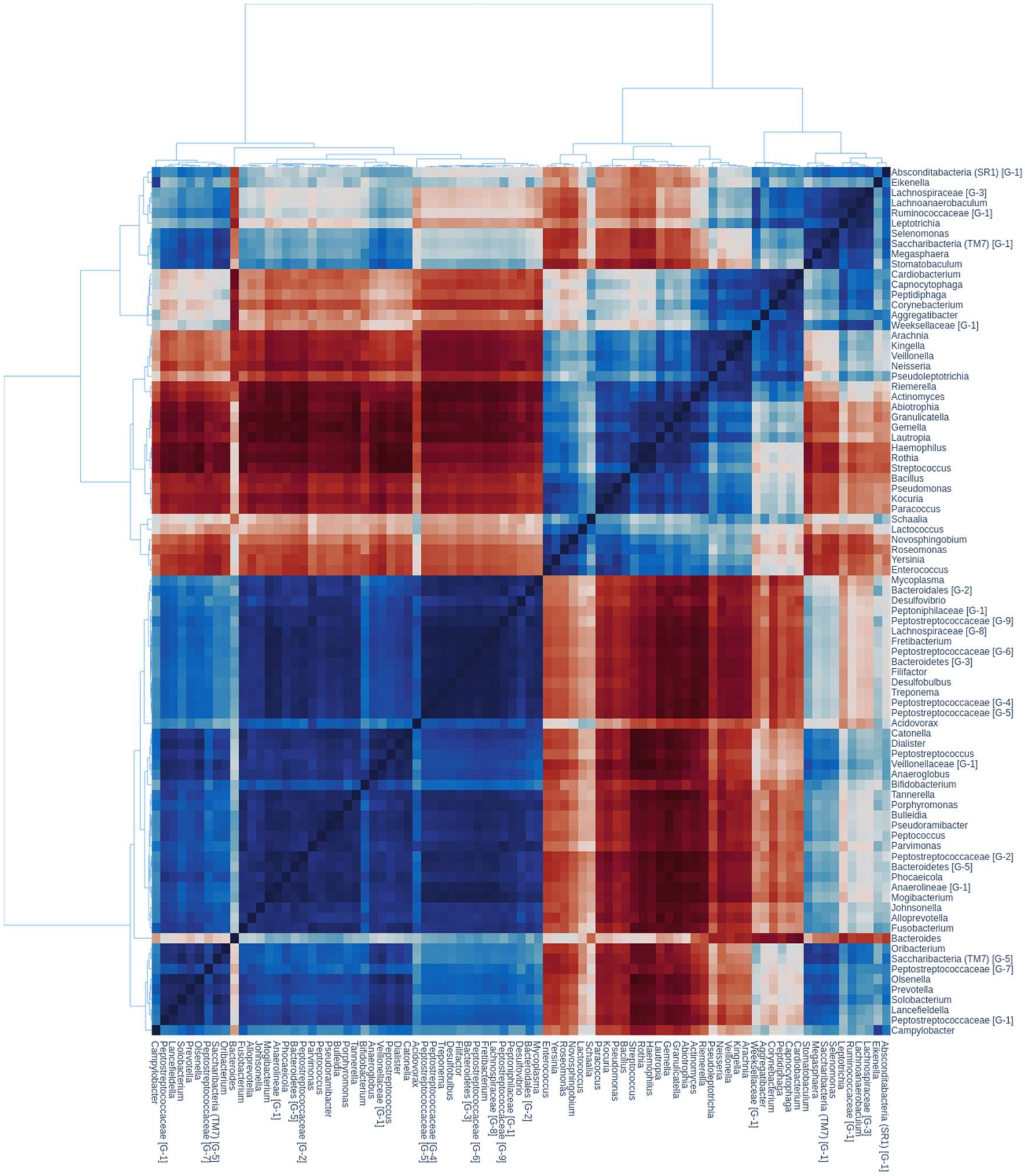

The heatmap displays the Spearman correlations of enrichment scores among microbiome genera. The genera are ordered based on hierarchical clustering of the Spearman correlations. The corresponding tree of hierarchical clustering is visualized next to the heatmap.

#### Figure S5 – Co-enrichment heatmap of non-microbiome phenotypes

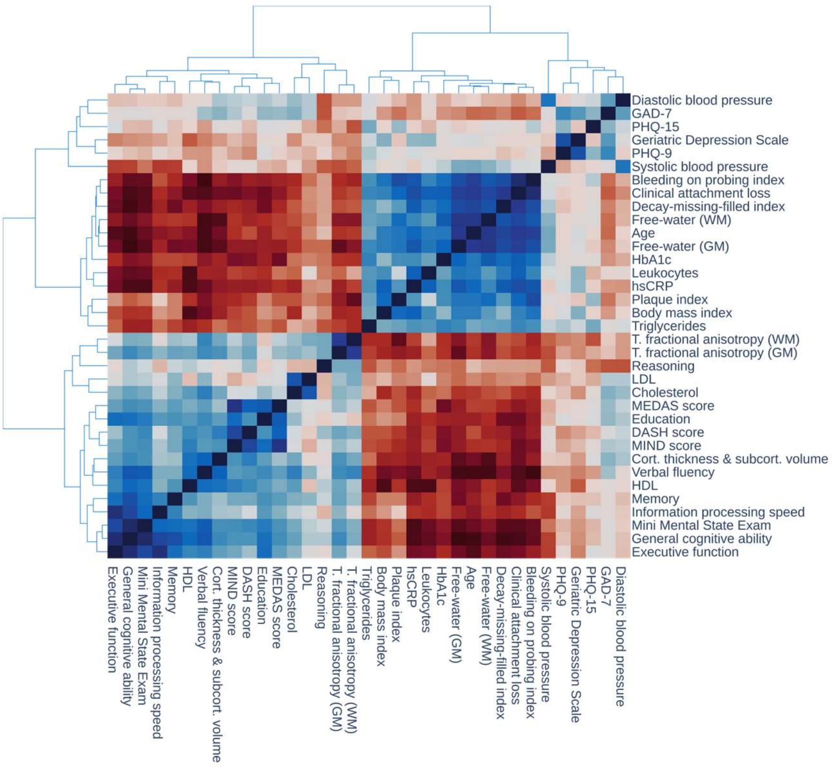

The heatmap displays the Spearman correlations of enrichment scores among microbiome genera. The genera are ordered based on hierarchical clustering of the Spearman correlations. The corresponding tree of hierarchical clustering is visualized next to the heatmap.

#### Figure S6 – Group comparison results for all bacterial genera

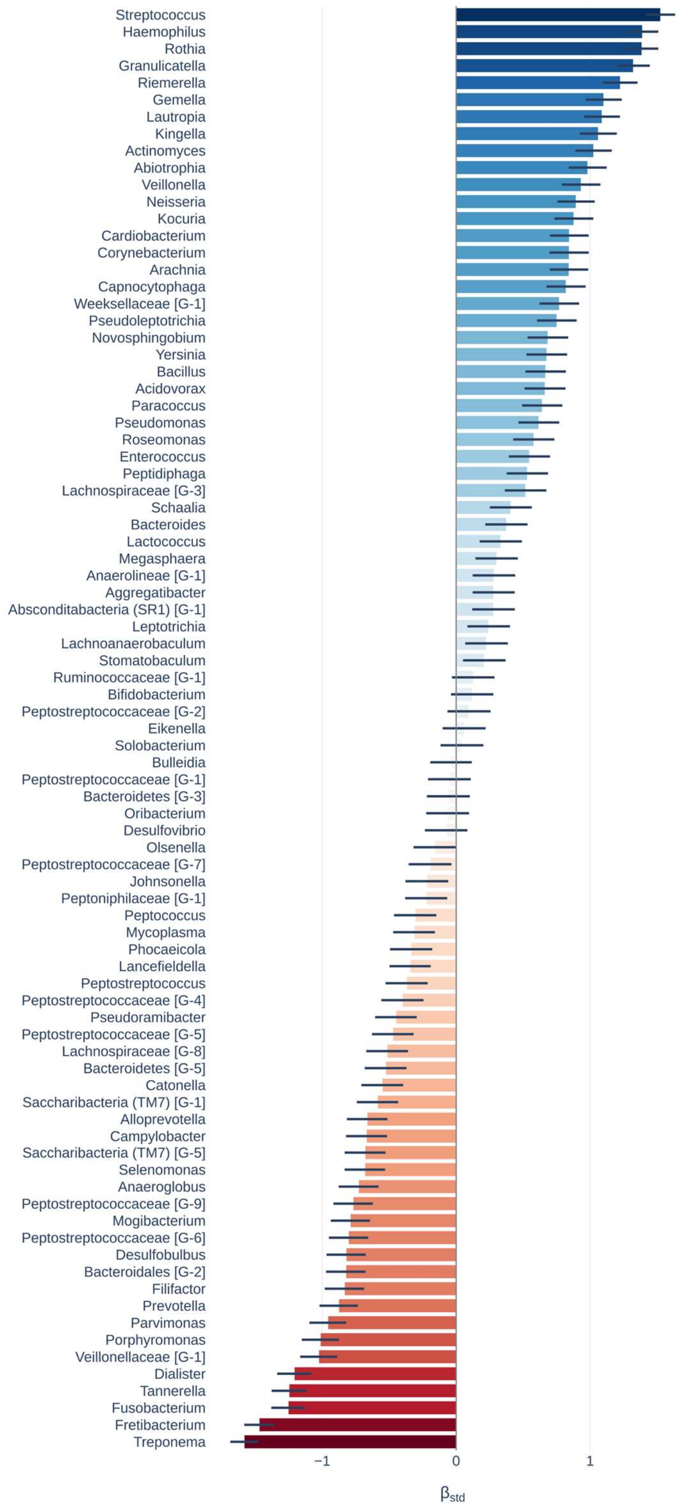

Barplot displays regression coefficients representing effects of group. Negative coefficients corresponding with higher abundance in the “A” group are shown in red, while positive coefficients indicating higher abundance in the “B” group are shown in blue. Error bars represent 95% confidence intervals.

#### Figure S7 – Group comparison results for non-significant clinical and lifestyle phenotypes

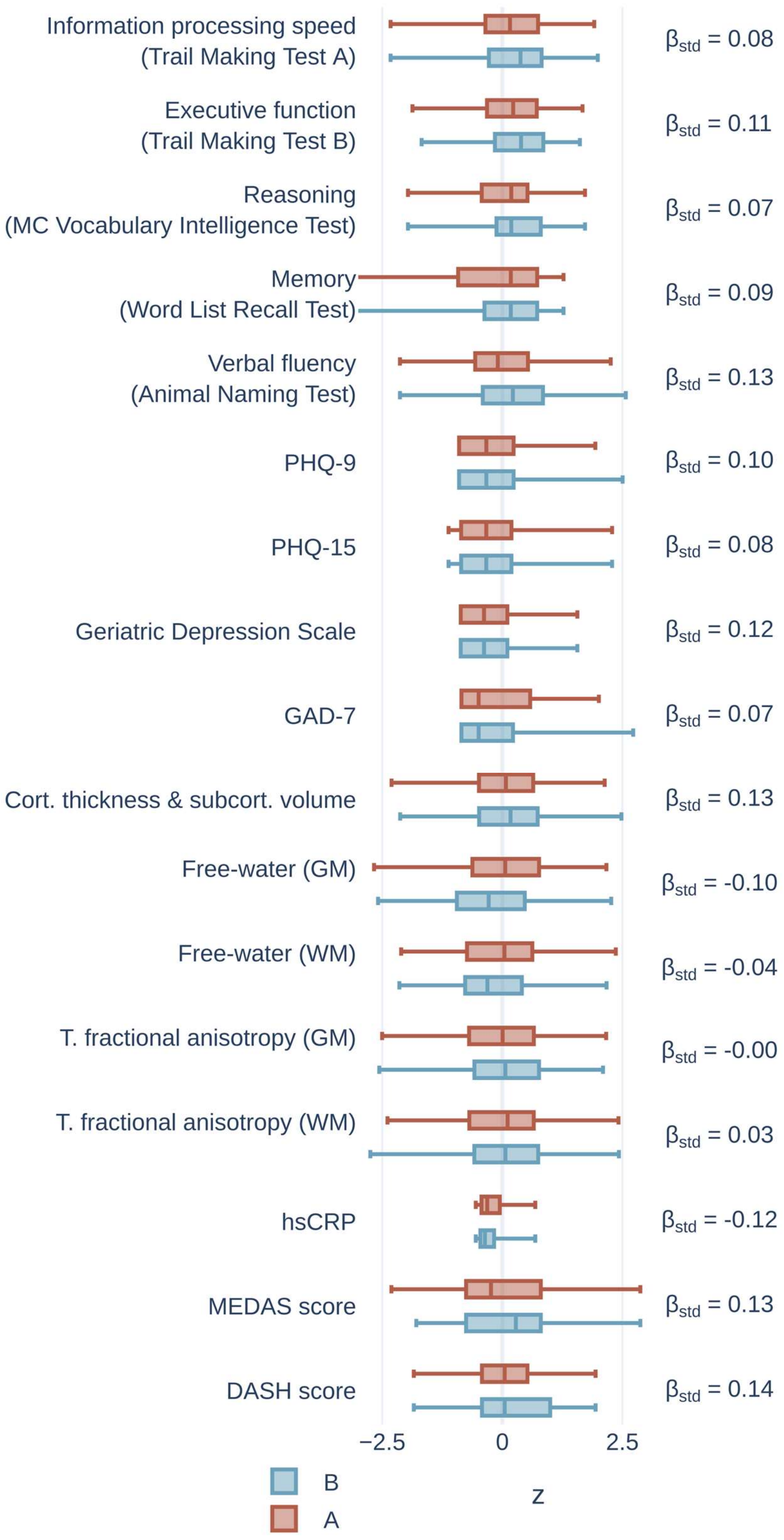

Box plot colors correspond to the groups.

#### Figure S8 – Confound analysis

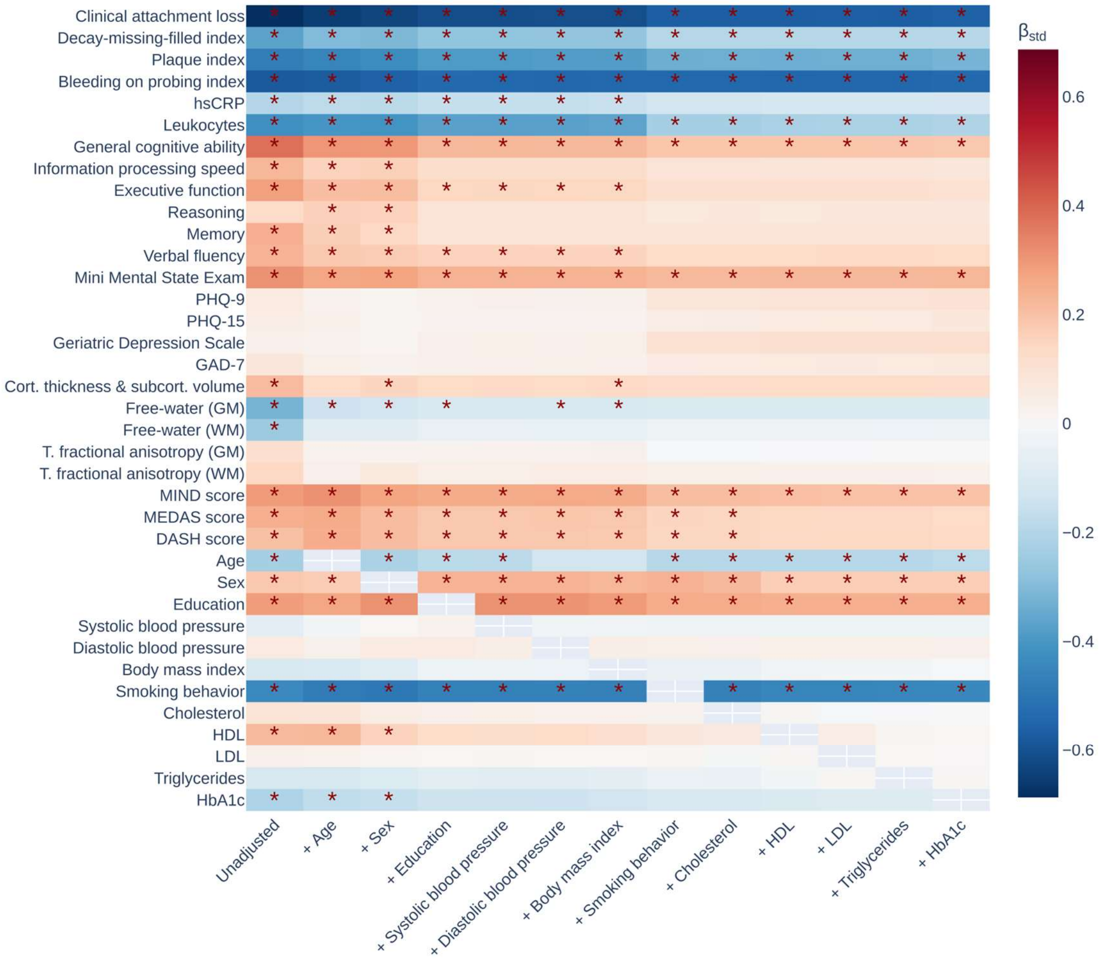

The heatmap illustrates the influence of sequential covariate adjustment on the association between microbiome group assignment and non-microbiome phenotypes. Each row represents a different phenotype, and columns represent the model at each step of adjustment, from unadjusted to fully adjusted. The color of each cell indicates the standardized beta coefficient ($\beta_{std}$) for the microbiome group variable, while red asterisks denote that the association remained statistically significant ($p < 0.05$).

#### Figure S9 – Sensitivity analysis across random subsamples of varying sample sizes

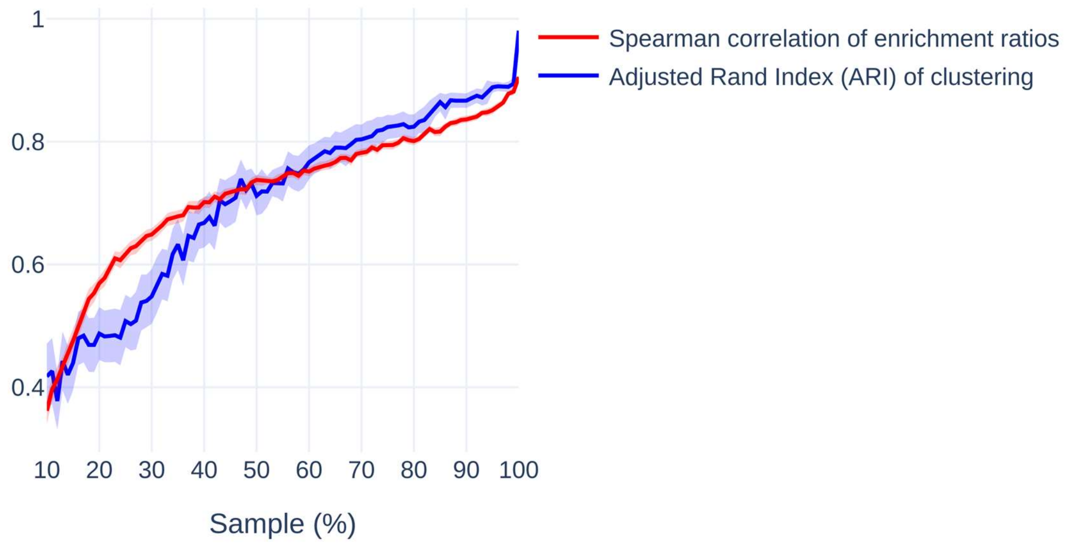

Line plot showing robustness indices as a function of sample size. The red curve represents the Spearman correlation coefficient between the enrichment ratios of non-microbiome and microbiome phenotypes in a randomly drawn subsample compared to the original enrichment ratios. The blue curve indicates the robustness of subject-to-group assignment using k-Means clustering before group analysis, measured by the Adjusted Rand Index. Displayed curves correspond with the average across 100 subsamples per %-step. Shaded areas denote 95% confidence intervals. Higher values on both measures indicate greater robustness.

#### Table S10 – Sensitivity analysis of pipeline design choices

| Pipeline design variation | Spearman correlation of enrichment ratio | Adjusted Rand Index of k-Means clustering |
|---|---|---|
| Mapper cover overlap: 1 [default: 1.5] | 0.80 | 0.81 |
| Mapper cover overlap: 1.2 [default: 1.5] | 0.82 | 0.81 |
| Mapper cover overlap: 1.4 [default: 1.5] | 0.81 | 0.88 |
| Mapper cover overlap: 1.6 [default: 1.5] | 0.79 | 0.91 |
| Mapper cover overlap: 1.8 [default: 1.5] | 0.84 | 0.91 |
| Mapper cover overlap: 2 [default: 1.5] | 0.83 | 0.90 |
| Mapper cover resolution: 20 [default: 30] | 0.88 | 0.72 |
| Mapper cover resolution: 25 [default: 30] | 0.85 | 0.81 |
| Mapper cover resolution: 35 [default: 30] | 0.82 | 0.87 |
| Mapper cover resolution: 40 [default: 30] | 0.85 | 0.88 |
| Mapper cover resolution: 45 [default: 30] | 0.82 | 0.87 |
| Mapper epsilon threshold for HDB-SCAN: 0.90 [default: 0.95] | 0.77 | 0.83 |
| Mapper epsilon threshold for HDB-SCAN: 0.99 [default: 0.95] | 0.83 | 0.76 |
| SAFE distance threshold: 0.5 [default: 0.75] | 0.85 | - * |
| SAFE distance threshold: 0.99 [default: 0.75] | 0.85 | - * |
| SAFE neighborhood radius: 0.05 [default: 0.1] | 0.84 | - * |
| SAFE neighborhood radius: 0.15 [default: 0.1] | 0.77 | - * |
| Mean ± SD | 0.82 ± 0.03 | 0.84 ± 0.06 |

* The pipeline adjustments involving SAFE do not alter the k-Means clustering of the topological network as they are not relevant for the computation.

Abbreviations: HDBSCAN = Hierarchical Density-Based Spatial Clustering of Applications with Noise, SAFE = Spatial Analysis of Functional Enrichment

#### Figure S11 – Computation of general cognitive ability (g) via principal component analysis

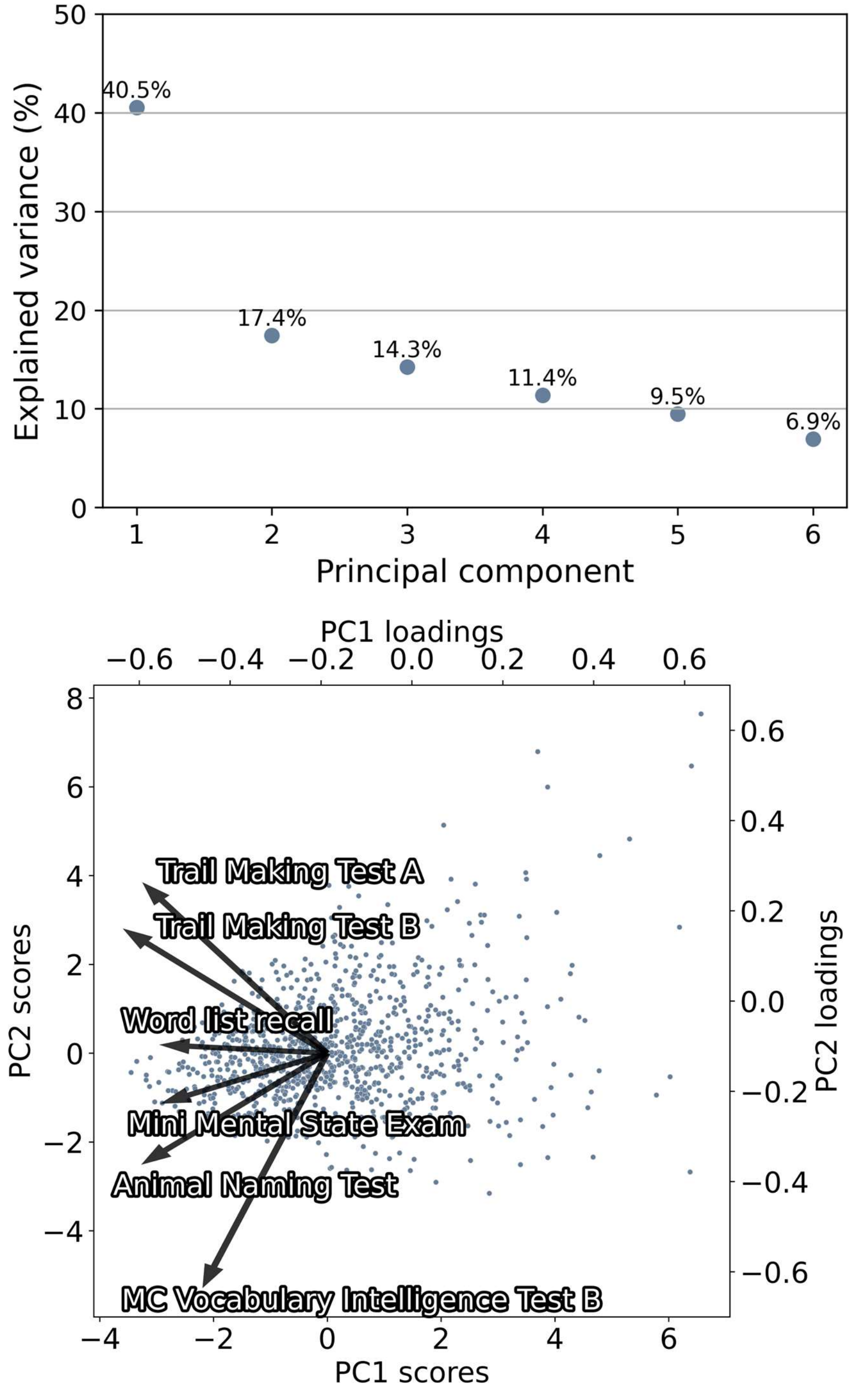

The scree plot displays the explained variance of derived principal components. The biplot displays the individual cognitive scores loadings on the principal components (arrows) and the scores of subjects (dots). *Abbreviations*: PC1 = principal component 1, PC2 = principal component 2.

#### Text S12 – Brain MRI acquisition, preprocessing and computation of neuroimaging markers

MR imaging was conducted using a single 3-Tesla Siemens Skyra. The scanning protocols follow those outlined in previous studies. For 3D T1-weighted anatomical images, a rapid acquisition gradient-echo sequence (MPRAGE) was used with the following sequence specifics: repetition time (TR) = 2500 ms, echo time (TE) = 2.12 ms, 256 axial slices, slice thickness (ST) = 0.94 mm, in-plane resolution (IPR) = 0.83 × 0.83 mm. 3D T2-weighted fluid-attenuated inversion recovery (FLAIR) images with TR = 4700 ms, TE = 392 ms, 192 axial slices, ST = 0.9 mm, and IPR = 0.75 × 0.75 mm. For single-shell diffusion-weighted imaging (DWI) 75 axial slices were acquired with gradients (b = 1000 s/mm$^2$) applied along 64 noncollinear directions with the following sequence parameters: TR = 8500 ms, TE = 75 ms, ST = 2 mm, IPR = 2 × 2 mm, anterior–posterior phase-encoding direction, 1 b0 volume.

Quality assessment included visual inspection of raw images and processed images based on quantitative outliers defined as measures exceeding 2 standard deviations from the mean of quality measures computed via QSIprep and mriqc.[2,3]

MRI data were processed employing preconfigured containerized pipelines as described previously.[4] The full documentation can be found on GitHub (https://github.com/csi-hamburg/CSIframe/wiki). Diffusion- and T1-weighted images were preprocessed employing QSIprep (v. 0.14.2).[2] DWI preprocessing included bias field correction, skull-stripping, denoising, deringing, as well as correction of B1 field inhomogeneity, head motion, eddy currents, and susceptibility distortions.[2,5–8] T1-weighted and FLAIR images underwent bias field correction and skull-stripping.[8]

Cortical thickness and volume. FreeSurfer (v. 6.0.1) was used to derive regional measures of cortical thickness and subcortical volumes.[9,10] After surface reconstruction, cortical thickness was measured as the distance between the white matter and pial surface.[10] Cortical thickness was averaged across the cortex. Subcortical volumes were averaged across all structures of the aseg atlas.[9] The mean cortical thickness and mean subcortical volume were z-scored. Subsequently, the two resulting z scores were averaged to obtain a single summary measure of cortical thickness and subcortical volume.

Free-water and tissue fractional anisotropy. Diffusion imaging features were derived from preprocessed diffusion-weighted images. We conducted free-water imaging, employing a dual-tensor model that delineates the isotropic extracellular compartment and the cellular compartment characterized by hindered or restricted diffusion. Based on a regularized non-linear fitting process, free-water and tissue fractional anisotropy ($FA_T$) were calculated.[11] Voxel-level free-water and tissue fractional anisotropy were averaged across gray matter voxels as well as across white matter voxels to obtain global measures for gray and white matter, respectively.
